# Deficient butyrate-producing capacity in the gut microbiome of Myalgic Encephalomyelitis/Chronic Fatigue Syndrome patients is associated with fatigue symptoms

**DOI:** 10.1101/2021.10.27.21265575

**Authors:** Cheng Guo, Xiaoyu Che, Thomas Briese, Orchid Allicock, Rachel A. Yates, Aaron Cheng, Amit Ranjan, Dana March, Mady Hornig, Anthony L. Komaroff, Susan Levine, Lucinda Bateman, Suzanne D. Vernon, Nancy G. Klimas, Jose G. Montoya, Daniel L. Peterson, W. Ian Lipkin, Brent L. Williams

## Abstract

**Background:** Myalgic Encephalomyelitis/Chronic Fatigue Syndrome (ME/CFS) is a complex, debilitating disease of unknown cause for which there is no specific therapy. Patients suffering from ME/CFS commonly experience persistent fatigue, post-exertional malaise, cognitive dysfunction, sleep disturbances, orthostatic intolerance, fever and irritable bowel syndrome (IBS). Recent evidence implicates gut microbiome dysbiosis in ME/CFS. However, most prior studies are limited by small sample size, differences in clinical criteria used to define cases, limited geographic sampling, reliance on bacterial culture or 16S rRNA gene sequencing, or insufficient consideration of confounding factors that may influence microbiome composition. In the present study, we evaluated the fecal microbiome in the largest prospective, case-control study to date (n=106 cases, n=91 healthy controls), involving subjects from geographically diverse communities across the United States.

**Results:** Using shotgun metagenomics and qPCR and rigorous statistical analyses that controlled for important covariates, we identified decreased relative abundance and quantity of *Faecalibacterium*, *Roseburia*, and *Eubacterium* species and increased bacterial load in feces of subjects with ME/CFS. These bacterial taxa play an important role in the production of butyrate, a multifunctional bacterial metabolite that promotes human health by regulating energy metabolism, inflammation, and intestinal barrier function. Functional metagenomic and qPCR analyses were consistent with a deficient microbial capacity to produce butyrate along the acetyl-CoA pathway in ME/CFS. Metabolomic analyses of short-chain fatty acids (SCFAs) confirmed that fecal butyrate concentration was significantly reduced in ME/CFS. Further, we found that the degree of deficiency in butyrate-producing bacteria correlated with fatigue symptom severity among ME/CFS subjects. Finally, we provide evidence that IBS comorbidity is an important covariate to consider in studies investigating the microbiome of ME/CFS subjects, as differences in microbiota alpha diversity, some bacterial taxa, and propionate were uniquely associated with self-reported IBS diagnosis.

**Conclusions:** Our findings indicate that there is a core deficit in the butyrate-producing capacity of the gut microbiome in ME/CFS subjects compared to healthy controls. The relationships we observed among symptom severity and these gut microbiome disturbances may be suggestive of a pathomechanistic linkage, however, additional research is warranted to establish any causal relationship. These findings provide support for clinical trials that explore the utility of dietary, probiotic and prebiotic interventions to boost colonic butyrate production in ME/CFS.

## Background

Myalgic Encephalomyelitis/Chronic Fatigue Syndrome (ME/CFS) is an unexplained debilitating and chronic disease characterized by a spectrum of symptoms including fatigue, post-exertional malaise, impaired memory, pain, gastrointestinal dysfunction, immune abnormalities, and sleep disturbances [1–3]. The global prevalence of ME/CFS ranges between 0.4 and 2.5%. The illness predominantly begins in adults 20 to 40 years of age, and is more common in females than males with a ratio averaging about 3:1, ranging as high as 6:1[4–7]. In the US alone this syndrome afflicts 2.5 million individuals [3], with an estimated economic burden of approximately $18 - $24 billion USD [8].

A diagnosis of ME/CFS is based on clinical criteria and symptoms utilizing different but overlapping case definitions including the 1994 U.S. Centers for Disease Control and Prevention (CDC), Canadian Consensus, International Consensus, and the Oxford and Institute of Medicine Criteria [1-3, 9, 10]. The cause or causes of ME/CFS are unknown. However, many underlying biological abnormalities have been identified in people with ME/CFS, including defective energy metabolism and a hypometabolic state; redox imbalance; dysregulated immune responses; multiple abnormalities of the central and autonomic nervous system; and multiple autoantibodies, many against targets in the central and autonomic nervous system, as summarized in two recent reviews [11, 12].

Many, but not all, people with ME/CFS report that their symptoms began following an acute prodromal viral-like syndrome [10]. Post-infection fatigue states are known to develop following viral, bacterial and protozoal infections [11]. Clusters of ME/CFS have also followed outbreaks of infectious disease [13, 14]. Interestingly, recent viral outbreaks including SARS-CoV, Ebola, MERS-CoV and SARS-CoV-2 have been associated with long-term sequelae that include persistent fatigue and cognitive symptoms consistent with those reported in ME/CFS [15–19].

The gut microbiome can influence human health and affect physiological systems through its influence on pathogen resistance, maintenance of the gut barrier, metabolism, immunity, and neural signaling. The gut microbiome directly affects immunity via the production of various immunomodulatory metabolites such as short-chain fatty acids (SCFAs), which support immunological tolerance and maintain inflammatory equilibrium [20–23]. Several studies, including our own, have identified differences in the gut microbiome (dysbiosis) between ME/CFS patients and healthy individuals [24–30]. However, most of these studies have limitations due to small sample size, insufficient assessment of potentially confounding variables, and biases associated with 16S sequencing for bacterial classification.

In this study, we employed fecal shotgun metagenomics and metabolomics to assess dysbiosis in the largest prospective case-control study to date, consisting of 106 ME/CFS subjects and 91 matched healthy controls. Our findings provide new insights into disturbances in the microbiota, functional metagenomic pathways, and SCFAs and their relationship with fatigue symptoms in ME/CFS.

## Results

### Study Population

The study included stool samples from 106 ME/CFS cases who met both the 1994 CDC and the 2003 Canadian consensus criteria for ME/CFS, and 91 healthy control individuals recruited from 5 sites in the United States. The characteristics of the ME/CFS patients and healthy participants of this study are outlined in **Table 1**. The majority of the ME/CFS cases were categorized as long duration, with 92.5% presenting with MECFS for longer than 3 years. Self-reported irritable bowel syndrome diagnosis (sr-IBS) was more frequent in ME/CFS cases (33.0%) compared to healthy controls (3.3%) (Fisher’s Exact test, p < 0.001), consistent with prior reports of higher IBS comorbidity in ME/CFS [31–33].

**Table 1:**
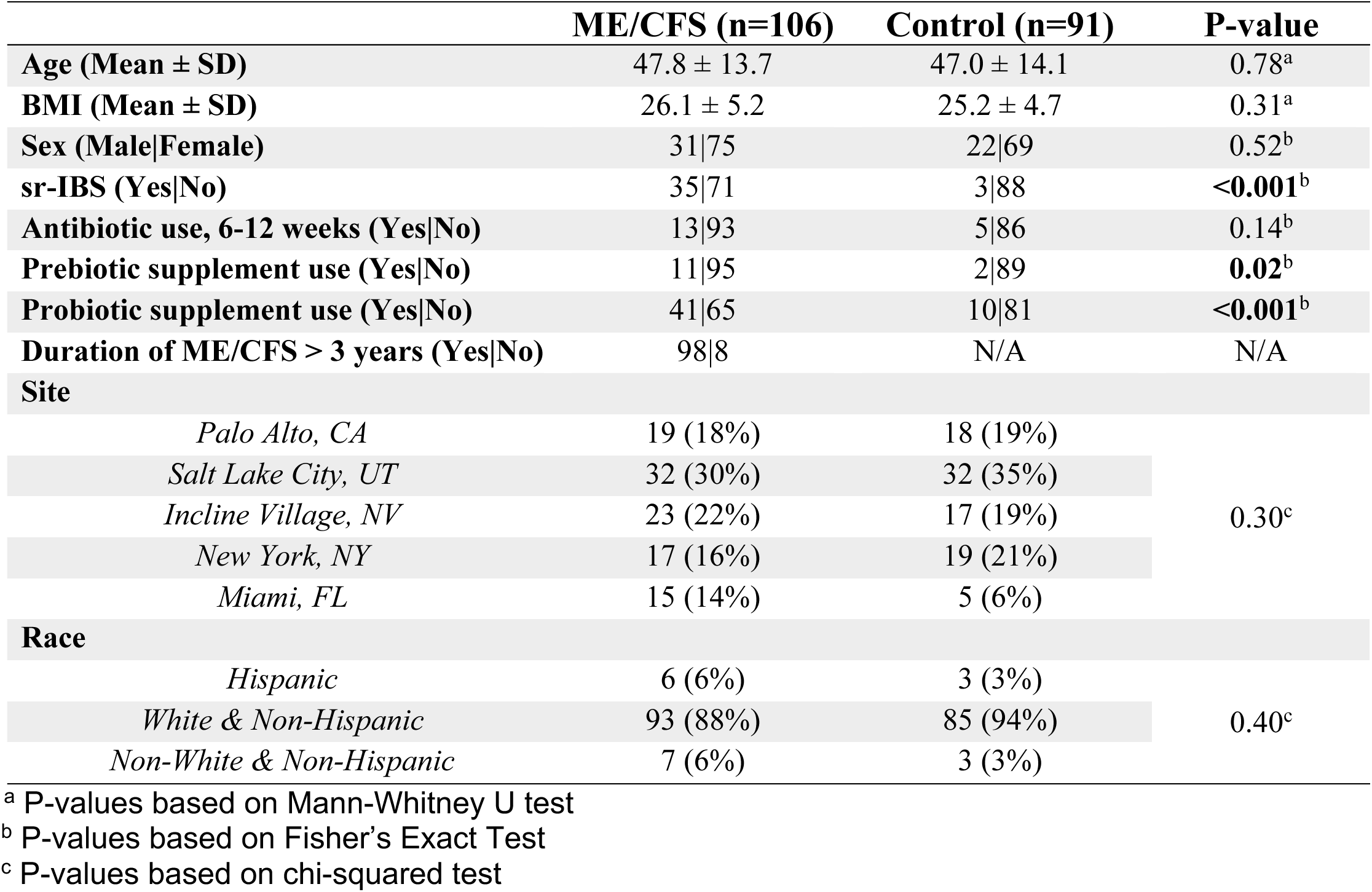
Cohort characteristics

|  | ME/CFS (n=106) | Control (n=91) | P-value |
| --- | --- | --- | --- |
| Age (Mean ± SD) | 47.8 ± 13.7 | 47.0 ± 14.1 | 0.78<sup>a</sup> |
| BMI (Mean ± SD) | 26.1 ± 5.2 | 25.2 ± 4.7 | 0.31<sup>a</sup> |
| Sex (Male\|Female) | 31\|75 | 22\|69 | 0.52<sup>b</sup> |
| sr-IBS (Yes\|No) | 35\|71 | 3\|88 | <0.001<sup>b</sup> |
| Antibiotic use, 6-12 weeks (Yes\|No) | 13\|93 | 5\|86 | 0.14<sup>b</sup> |
| Prebiotic supplement use (Yes\|No) | 11\|95 | 2\|89 | 0.02<sup>b</sup> |
| Probiotic supplement use (Yes\|No) | 41\|65 | 10\|81 | <0.001<sup>b</sup> |
| Duration of ME/CFS > 3 years (Yes\|No) | 98\|8 | N/A | N/A |
| Site |  |  |  |
| Palo Alto, CA | 19 (18%) | 18 (19%) | 0.30<sup>c</sup> |
| Salt Lake City, UT | 32 (30%) | 32 (35%) |  |
| Incline Village, NV | 23 (22%) | 17 (19%) |  |
| New York, NY | 17 (16%) | 19 (21%) |  |
| Miami, FL | 15 (14%) | 5 (6%) |  |
| Race |  |  |  |
| Hispanic | 6 (6%) | 3 (3%) | 0.40<sup>c</sup> |
| White & Non-Hispanic | 93 (88%) | 85 (94%) |  |
| Non-White & Non-Hispanic | 7 (6%) | 3 (3%) |  |

<sup>a</sup> P-values based on Mann-Whitney U test
<sup>b</sup> P-values based on Fisher's Exact Test
<sup>c</sup> P-values based on chi-squared test

The site of collection, age, sex, BMI, and demographic index were matched in healthy controls. More ME/CFS females enrolled in the study (70.8% of all ME/CFS cases), consistent with a higher prevalence of ME/CFS in females in the general population. Healthy controls consisted of 75.8% females. Mean age was similar between ME/CFS and healthy controls, 47.8 vs 47.0 years, respectively. Mean BMI was also similar between cases and controls, and 50% of the cases and 52.7% of the controls had a high BMI (over 25kg/m^2^).

No cases or controls took an antibiotic in the six weeks prior to stool sample collection (see participant exclusion criteria), and antibiotic use in the 6-12 week period prior to sample collection was similar between cases and controls. As prebiotic and probiotic supplement use can influence the microbiome, we compared usage between cases and controls. More ME/CFS subjects reported frequent prebiotic (Fisher’s Exact test, p = 0.02) and probiotic (Fisher’s Exact test, p < 0.001) supplement usage compared to healthy controls.

### Characterization of the gut microbiome

Shotgun metagenomic sequencing of stool samples from all 106 ME/CFS and 91 healthy controls was undertaken to assess bacterial composition, alpha and beta diversity, differential abundance of bacterial taxa, and predicted functional metabolic pathways. On average, >27 million (range: 12-68 million) raw sequences were obtained per sample. After quality filtering and host subtraction, bacterial reads were classified using Kraken2 and relative abundances of bacteria were estimated using Bracken. At the genus-level, across the dataset, *Bacteroides* was the most abundant genus in both ME/CFS cases and healthy controls, consistent with prior reports of *Bacteroides* dominance in individuals in the United States (**Figure 1A**) [34]. Other high abundance genera observed in ME/CFS and healthy controls included *Alistipes*, *Parabacteroides*, *Roseburia*, *Prevotella*, and *Faecalibacterium*.

**Figure 1:**
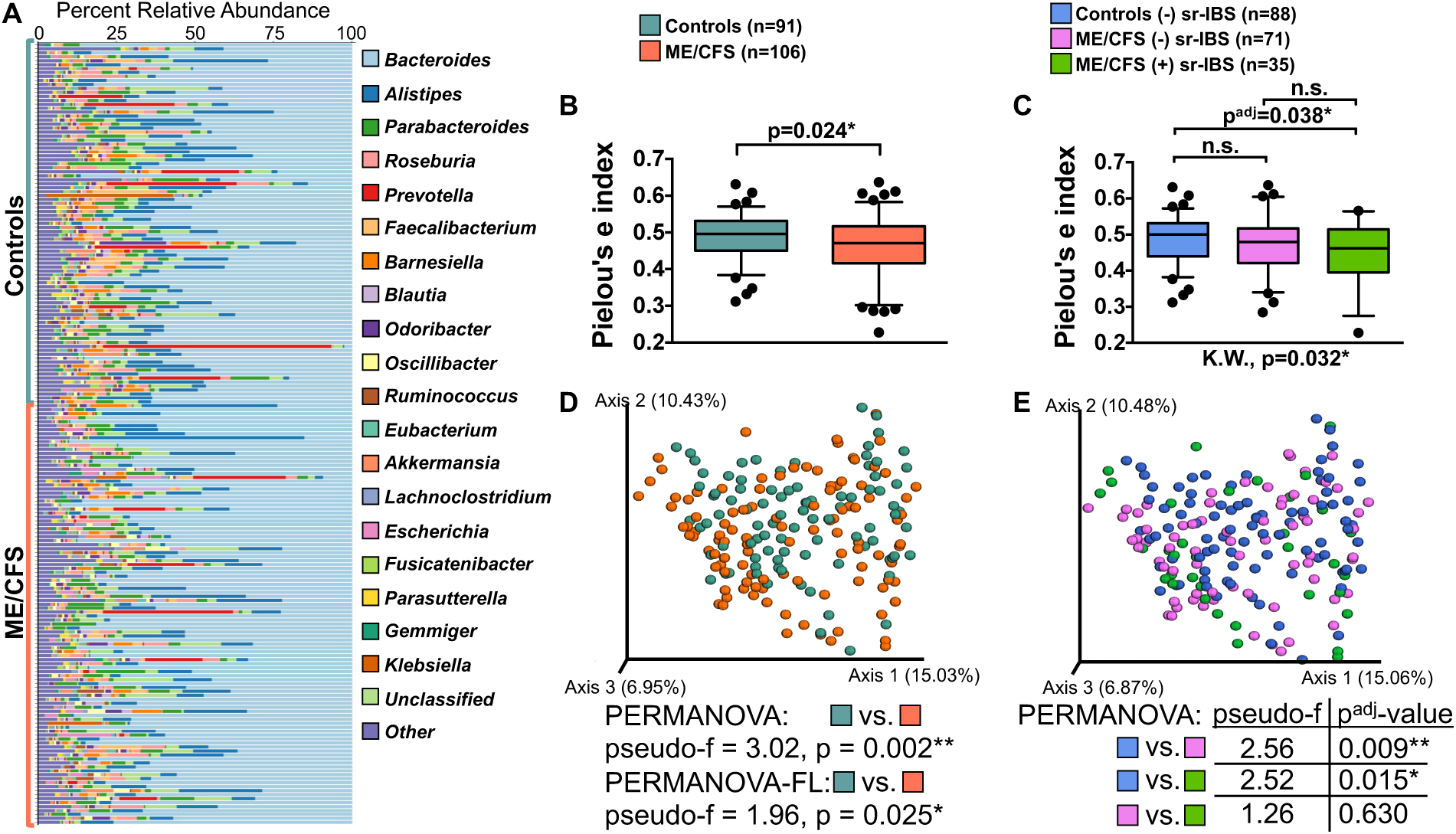
Gut microbiome diversity. Stacked bar chart showing the percent relative abundance of the most abundant bacterial genera in ME/CFS (n=106) and healthy control subjects (n=91) (**A**). Box-and-whiskers plots showing the distribution of microbiome alpha diversity based on Pielou’s e index (evenness) between ME/CFS and healthy controls (**B**) and among stratified groups for healthy controls without (-) sr-IBS (n=88), ME/CFS subjects without (-) sr-IBS (n=71), and ME/CFS subjects with (+) sr-IBS (n=35) (**C**). Statistical significance was determined based on two-tailed p-values from the Mann-Whitney U test (**B**). For stratified analyses significance was first determined based on the Kruskal-Wallis test (K.W., results shown below figure in **C**) and between group significance was determined based on the Mann-Whitney U test with multiple testing (Bonferroni) correction (p^adj^-value). n.s. = not significant; * = p or p^adj^ < 0.05; ** = p or p^adj^ < 0.01. PCoA plots of microbiota beta diversity based on the Bray-Curtis dissimilarity metric comparing ME/CFS and healthy controls (**D**) and groups stratified by sr-IBS status (**E**). Differences based on PERMANOVA (unadjusted) (**D**, **E**) or PERMANOVA-FL (adjusted for covariates) (**D**) are shown below for each group comparison with associated pseudo-f statistic and p-value (**D**) or the Bonferroni adjusted p-value (p^adj^) (**E**).

Bacterial alpha diversity metrics (Pielou’s evenness, Shannon index, and observed species) were compared in univariate analyses to assess differences in alpha diversity between ME/CFS cases and healthy controls. The microbiota of ME/CFS patients had lower evenness (Mann-Whitney U test, p = 0.024) and Shannon diversity (Mann-Whitney U test, p = 0.023) (**Figure 1B** and **Supplementary Figure 1A**) but similar observed species (**Supplementary Figure 1B**) compared to healthy controls.

IBS has also been associated with changes in the gut microbiota, including alpha diversity [35–38]. Given that the frequency of sr-IBS diagnosis was higher in ME/CFS in this study, and that our prior research indicated that comorbid sr-IBS may be associated with alterations in microbiota in ME/CFS [25], we performed stratified analyses to assess differences in alpha diversity among healthy controls without sr-IBS (n=88), ME/CFS without sr-IBS (n=71) and ME/CFS with sr-IBS (n=35). As only three healthy controls had sr-IBS diagnosis in our cohort, no stratified comparisons with this group were undertaken. In stratified analyses, evenness and Shannon diversities were only lower in ME/CFS with sr-IBS compared with healthy controls without sr-IBS (Mann-Whitney U test, p^adj^ = 0.038 and p^adj^ = 0.047, respectively) (**Figure 1C** and **Supplementary Figure 1C**). No differences were found between stratified groups for observed species (**Supplementary Figure 1D**). These results indicate that differences in alpha diversity between ME/CFS and controls may be dependent on comorbid sr-IBS, rather than ME/CFS.

To further evaluate the relationship between alpha diversity and ME/CFS, we employed linear regression (Shannon and evenness) and negative binomial regression (observed species) with alpha diversity metrics as outcome variables and ME/CFS and sr-IBS as predictors, adjusting for covariates of site of sampling, sex, BMI, race/ethnicity, age, antibiotic usage 6-12 weeks prior to sample collection, probiotic supplement use, and prebiotic supplement use (**Table 2**). The interaction term between ME/CFS and sr-IBS was explored but was not included in the final model as it was not significant in any of the regression models for alpha diversity. ME/CFS was not a significant predictor of evenness, Shannon diversity, or observed species metrics. However, sr-IBS status was negatively associated with evenness (β_Est_ = -0.03, 95% CI: -0.06-0.00, p = 0.032) and showed a trending negative association with Shannon diversity ((β_Est_ = -0.03, 95% CI: - 0.06-0.00, p = 0.054). Of other covariates, only subject age showed a significant positive association with all three alpha diversity metrics. These results suggest that differences in alpha diversity are associated with sr-IBS rather than ME/CFS and highlight the importance of assessing IBS as a potential confounder of microbiota differences in ME/CFS.

**Table 2:** Generalized linear regression showing the relationship between ME/CFS status and alpha diversity metrics.

| Predictors | *shannon* |  |  | *evenness* |  |  | *observed species* |  |  |
| --- | --- | --- | --- | --- | --- | --- | --- | --- | --- |
| | $\beta_{Est}$ | 95% CI | p | $\beta_{Est}$ | 95% CI | p | Fold Change | 95% CI | p |
| (Intercept) | 0.63 | 0.60 - 0.67 | **<0.001** | -0.31 | -0.34 - -0.28 | **<0.001** | 432.26 | 400.00 - 467.11 | **<0.001** |
| ME/CFS | -0.02 | -0.05 - 0.00 | 0.103 | -0.02 | -0.04 - 0.00 | 0.096 | 0.98 | 0.93 - 1.03 | 0.481 |
| sr-IBS | -0.03 | -0.06 - 0.00 | 0.054 | -0.03 | -0.06 - 0.00 | **0.032** | 1.01 | 0.94 - 1.07 | 0.865 |
| Site 2(Florida) | 0.00 | -0.05 - 0.05 | 0.973 | 0.00 | -0.04 - 0.04 | 0.954 | 0.99 | 0.90 - 1.09 | 0.859 |
| Site 3(Nevada) | -0.02 | -0.06 - 0.02 | 0.246 | -0.02 | -0.05 - 0.02 | 0.279 | 0.95 | 0.88 - 1.03 | 0.222 |
| Site 4(New York) | 0.01 | -0.03 - 0.04 | 0.673 | 0.00 | -0.03 - 0.04 | 0.814 | 1.05 | 0.98 - 1.14 | 0.166 |
| Site 5(Utah) | 0.00 | -0.03 - 0.04 | 0.863 | 0.00 | -0.03 - 0.03 | 0.957 | 1.02 | 0.95 - 1.10 | 0.522 |
| Sex | -0.01 | -0.04 - 0.02 | 0.582 | -0.01 | -0.03 - 0.02 | 0.669 | 0.97 | 0.91 - 1.03 | 0.268 |
| BMI | 0.00 | -0.01 - 0.01 | 0.920 | 0.00 | -0.01 - 0.01 | 0.726 | 0.98 | 0.96 - 1.00 | 0.119 |
| Hispanic | 0.01 | -0.05 - 0.06 | 0.850 | 0.00 | -0.05 - 0.06 | 0.886 | 1.02 | 0.90 - 1.15 | 0.758 |
| Non-White & Non-Hispanic | -0.02 | -0.07 - 0.03 | 0.370 | -0.02 | -0.07 - 0.03 | 0.395 | 0.94 | 0.85 - 1.05 | 0.291 |
| Age | 0.02 | 0.00 - 0.03 | **0.016** | 0.01 | 0.00 - 0.02 | **0.023** | 1.03 | 1.01 - 1.06 | **0.011** |
| Antibiotics | 0.01 | -0.03 - 0.05 | 0.677 | 0.01 | -0.02 - 0.05 | 0.520 | 0.96 | 0.88 - 1.04 | 0.292 |
| Probiotics | 0.02 | -0.01 - 0.05 | 0.237 | 0.02 | -0.01 - 0.04 | 0.186 | 1.00 | 0.94 - 1.06 | 0.871 |
| Prebiotics | -0.04 | -0.09 - 0.01 | 0.125 | -0.03 | -0.08 - 0.01 | 0.133 | 0.94 | 0.85 - 1.04 | 0.223 |

Results for each outcome variable were estimated using linear regression (Shannon and evenness) or a negative binomial generalized linear model (observed species) and adjusted for covariates.
$\beta_{Est}$ (Estimate) or Fold Change, 95% Confidence Intervals (CI), and p-values are shown for each model.
Significant predictors (p<0.05) are indicated in bold.

Differences in microbiota beta diversity based on the Bray-Curtis dissimilarity between ME/CFS and healthy controls were assessed by PERMANOVA (unadjusted) and PERMANOVA-FL (adjusted for sr-IBS status, site of sampling, sex, BMI, race/ethnicity, age, antibiotic usage 6-12 weeks prior to sample collection, probiotic supplement use, and prebiotic supplement use). Although PCoA plots did not show clear separation of cases and controls, ME/CFS subjects differed from healthy controls based on Bray-Curtis dissimilarity even after adjusting for covariates (PERMANOVA, p = 0.002; PERMANOVA-FL, p = 0.025) (**Figure 1D**). We performed stratified analyses based on sr-IBS diagnosis to assess the possibility that differences in microbiota beta diversity were driven by sr-IBS diagnosis (**Figure 1E**). Unlike alpha diversity, beta diversity differed between healthy controls without sr-IBS and both ME/CFS without sr-IBS (PERMANOVA, p^adj^ =0.009) and ME/CFS with sr-IBS (PERMANOVA, p^adj^ =0.015). Beta diversity did not differ between ME/CFS without and with sr-IBS. Thus, beta diversity differences between ME/CFS and healthy controls are independent of sr-IBS diagnosis.

Our results suggest that whereas some differences in microbiota between ME/CFS and healthy controls may be confounded by sr-IBS diagnosis, other differences may be independent of sr-IBS and specific to ME/CFS. Thus, we employed both unstratified and stratified analyses and include sr-IBS diagnosis as a covariate in all regression models.

### Differential abundance of bacterial taxa

We employed Generalized Additive Models for Location, Scale and Shape with a zero-inflated beta distribution (GAMLSS-BEZI) to evaluate bacterial taxa that differ in relative abundance in ME/CFS [39]. To identify differentially abundant species that are unique to ME/CFS as well as those that are unique to ME/CFS with sr-IBS, we compared four models. Model 1 represented the unstratified comparison of all ME/CFS vs. healthy controls. Model 2 represented the stratified comparison of ME/CFS without sr-IBS vs. healthy controls without sr-IBS. Model 3 represented the stratified comparison of ME/CFS with sr-IBS vs. healthy controls without sr-IBS. Model 4 represented the stratified comparison of ME/CFS with sr-IBS vs. ME/CFS without sr-IBS (note; Model 4 did not produce any significant taxa differences and is therefore not shown). For each of Models 1-3 ME/CFS status was the variable for comparison, while sr-IBS status was the variable for comparison in Model 4, and each model is adjusted for covariates.

At the species-level, Model 1 identified four species after adjusting for multiple comparisons that differentiated ME/CFS from healthy controls. Model 2 identified two species that differentiated ME/CFS without sr-IBS from healthy controls without sr-IBS. Model 3 identified fifteen species that differentiated ME/CFS with sr-IBS from healthy controls without sr-IBS (**Figure 2A** and **Supplementary Table 1A**). The relative abundance of two species, *Eubacterium rectale* and *Faecalibacterium prausnitzii* were lower in the ME/CFS group in all three models. These results suggest that deficiency of these species in ME/CFS are independent of sr-IBS and other covariates. The most differentially abundant species were found in Model 3, where fifteen species were associated with ME/CFS with sr-IBS compared to healthy controls without sr-IBS. The majority of identified species (11/15) in Model 3 showed no overlap with other models suggesting that sr-IBS has an independent association with changes in the microbiota. The species associated with ME/CFS with sr-IBS included decreased relative abundance of *Alistipes putredinis*, *Dorea longicatena*, *Odoribacter splanchnicus*, and *Lachnospiraceae bacterium GAM79*, among others, and increased relative abundance of *Clostridium bolteae* and *Flavonifractor plautii*. It was also apparent from our analysis of Model 1 that several bacterial species from the same genera such as *Eubacterium* and *Roseburia* were significantly associated with ME/CFS before FDR adjustment (**Supplementary Table 1A**). Accordingly, we performed the same GAMLSS-BEZI analyses at the genus level.

**Figure 2:**
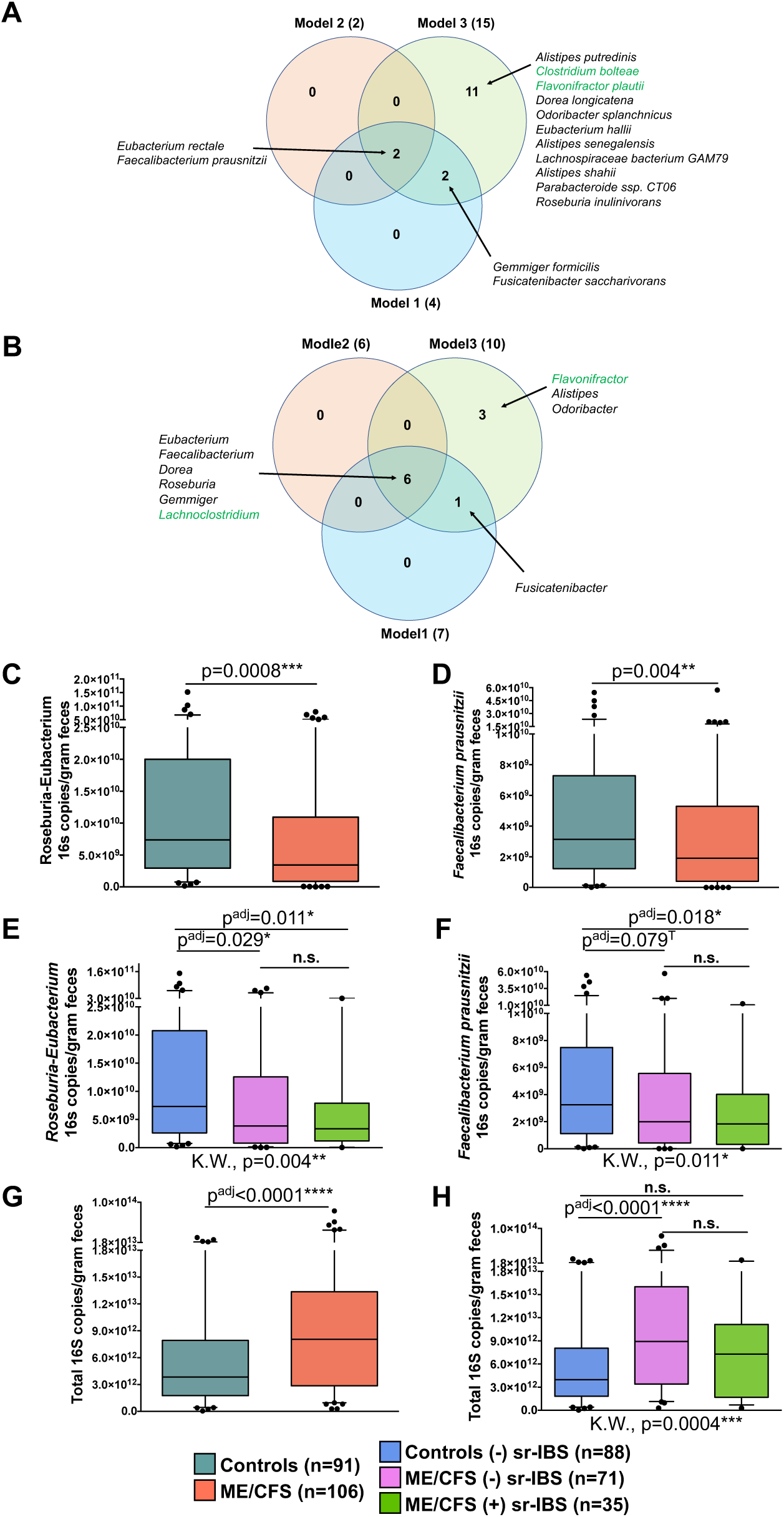
Differential abundance and quantitation of fecal bacteria. Venn diagram showing the results of multivariate GAMLSS-BEZI models for differentially abundant species (**A**) and genera (**B**) from four models: Model 1: comparing All ME/CFS vs healthy controls, Model 2: comparing ME/CFS without sr-IBS vs healthy controls without sr-IBS, Model 3: comparing ME/CFS with sr-IBS vs healthy controls without sr-IBS, and Model 4: comparing ME/CFS without sr-IBS vs ME/CFS with sr-IBS. Models are adjusted for covariates: sr-IBS diagnosis (only model 1), site of recruitment, sex, BMI, race and ethnicity, age, antibiotic use within 6-12 weeks of sample collection, probiotic supplement use, and prebiotic supplement use. Note: no taxa reached significance in Model 4 after FDR adjustment, thus only Models 1-3 are shown. Also see **Supplementary Tables 1A, B**. Box-and-whiskers plots showing the distribution of *Roseburia-Eubacterium* 16S rRNA and *F. prausnitzii* 16S rRNA per gram of feces determined by qPCR between ME/CFS and healthy controls (**C, D**) and among stratified groups for healthy controls without (-) sr-IBS, ME/CFS subjects without (-) sr-IBS, and ME/CFS subjects with (+) sr-IBS (**E, F**). Box-and-whiskers plots showing the distribution of Total bacterial 16S rRNA genes per gram of feces determined by qPCR between ME/CFS and healthy controls (**G**) and among stratified groups (**H**). Statistical significance was determined based on two-tailed p-values from the Mann-Whitney U test (**C, D, G**). For stratified analyses significance was first determined based on the Kruskal-Wallis test (K.W., results shown below each figure in **E, F, H**). If significant (p < 0.05) based on K.W., then between group significance was determined based on the Mann-Whitney U test with multiple testing (Bonferroni) correction (p^adj^-value). n.s. = not significant; * = p or p^adj^ < 0.05; ** = p or p^adj^ < 0.01; *** = p or p^adj^ < 0.001; T = trend (p or p^adj^ < 0.1).

At the genus-level six bacterial genera were significantly associated with ME/CFS from all three models after FDR adjustment (**Figure 2B** and **Supplementary Table 1B**). The majority of these genera had lower relative abundance in the ME/CFS group compared to healthy controls, including *Eubacterium*, *Faecalibacterium*, *Dorea*, *Roseburia* and *Gemmiger*. Only *Lachnoclostridium* was increased in relative abundance in the ME/CFS groups.

At both the species- and genus-level the primary bacteria identified in all three models, and thus associated with ME/CFS independent of sr-IBS, contained the most abundant and common butyrate-producing bacteria (BPB) in the human gut, including the species *E. rectale*, *F. prausnitzii* and the genera *Eubacterium*, *Faecalibacterium* and *Roseburia*.

### Quantitation of BPB in fecal samples

While shotgun metagenomics can provide compositional information on individual fecal bacteria, relative abundance of each taxon is dependent on the relative abundance of all bacterial taxa in the microbiota and is not strictly quantitative. In order to assess quantitative differences in fecal BPB, we carried out qPCR analysis using assays targeting the 16S rRNA genes of *Roseburia*-*Eubacterium* genera and the species *F. prausnitzii* to evaluate the quantity of these bacterial taxa per gram of feces. Compared to healthy controls, ME/CFS subjects had significantly fewer 16S rRNA gene copies of *Roseburia*-*Eubacterium* genera (Mann-Whitney U test, p = 0.0008) (**Figure 2C**) and the species *F. prausnitzii* (Mann-Whitney U test, p = 0.004) (**Figure 2D**) per gram of feces. In stratified analyses, *Roseburia*-*Eubacterium* 16S copies/gram of feces were lower in both ME/CFS subjects without sr-IBS (Mann-Whitney U test, p^adj^ = 0.029) and ME/CFS subjects with sr-IBS (Mann-Whitney U test, p^adj^ = 0.011) compared to healthy controls without sr-IBS (**Figure 2E**). Similarly, lower quantities of *F. prausnitzii* 16S/gram of feces were found when comparing ME/CFS without sr-IBS (Mann-Whitney U test, p^adj^ = 0.079) and ME/CFS with sr-IBS (Mann-Whitney U test, p^adj^ = 0.018) compared with healthy controls without sr-IBS, though only a trend was observed in the former after adjusting for multiple comparisons (**Figure 2F**).

As total bacterial load in stool has never been evaluated in ME/CFS, we carried out qPCR using a broad range 16S rRNA gene-targeting assay. In contrast to the quantitatively measured deficiencies in BPB in ME/CFS, ME/CFS subjects had higher quantities of total 16S copies/gram of feces compared to healthy controls (Mann-Whitney U test, p < 0.0001) (**Figure 2G**). Stratified analyses revealed that ME/CFS subjects without sr-IBS had higher quantities of total bacterial 16S/gram of feces compared with healthy controls without sr-IBS (Mann-Whitney U test, p^adj^ < 0.0001), while other group comparisons did not differ after adjusting for multiple comparisons (**Figure 2H**).

Since we measured both the quantity of BPB 16S and total bacterial 16S, we also calculated the relative abundance of *Roseburia*-*Eubacterium* and *F. prausnitzii* (i.e., *Roseburia*-*Eubacterium* 16S/total bacterial 16S). For both *Roseburia*-*Eubacterium* and *F. prausnitzii*, ME/CFS subjects had lower calculated relative abundance compared with healthy controls (Mann-Whitney U test, p = 0.0007 and p = 0.009, respectively) (**Supplementary Figure 2A, B**). In stratified analyses, *Roseburia*-*Eubacterium* calculated relative abundance was lower in ME/CFS subjects without sr-IBS compared with healthy controls without sr-IBS (Mann-Whitney U test, p^adj^ = 0.004), while other comparisons did not reach significance after adjusting for multiple comparisons (**Supplementary Figure 2C**). Similar results were obtained for *F. prausnitzii* calculated relative abundance, although only a trend toward lower *F. prausnitzii* was observed comparing ME/CFS subjects without sr-IBS with healthy controls without sr-IBS (Mann-Whitney U test, p^adj^ = 0.054) (**Supplementary Figure 2D**).

To further assess the relationships between quantitative measures of BPB and total bacteria (outcome variables) in feces with ME/CFS status (main predictor), generalized linear regression models with a Gamma distribution with log link were fit adjusting for covariates (**Table 3**). The estimated coefficients are interpreted as the log of fold-change (FC). ME/CFS status was significantly associated with lower quantities of *Roseburia-Eubacterium* 16S/gram of feces (FC=0.56, 95% CI: 0.33-0.93, p = 0.026) and *F. prausnitzii* 16S/gram of feces (FC=0.61, 95% CI: 0.38-0.98, p = 0.043) and higher quantities of total bacterial 16S/gram of feces (FC=2.08, 95% CI: 1.52-2.83). No other covariates were associated with *Roseburia*-*Eubacterium* quantity in the regression model, while BMI and antibiotic use were associated with *F. prausnitzii* quantity and BMI, antibiotic use and site of patient recruitment (Florida) were associated with total bacterial 16S. Similar results were obtained in regression models assessing the measured relative abundance of *Roseburia*-*Eubacterium* (FC=0.62, 95% CI: 0.42-0.92, p = 0.019) and *F. prausnitzii* (FC=0.60, 95% CI: 0.42-0.85, p = 0.004) (**Supplementary Table 2**).

**Table 3:** Generalized linear regression showing the relationship between ME/CFS status and fecal quantities of bacterial taxa and the *but* gene by qPCR.

| Predictors | *Roseburia/Eubacterium* 16S |  |  | *F. prausnitzii* 16S |  |  | total 16S |  |  | *but* gene |  |  |
| --- | --- | --- | --- | --- | --- | --- | --- | --- | --- | --- | --- | --- |
|  | Fold Change | 95% CI | p | Fold Change | 95% CI | p | Fold Change | 95% CI | p | Fold Change | 95% CI | p |
| (Intercept) | 1E+10 | 8E+9-3E+10 | **<0.001** | 7.03E+9 | 3E+9-1E+10 | **<0.001** | 4E+12 | 3E+12-7E+12 | **<0.001** | 8E+8 | 3E+8-2E+09 | **<0.001** |
| ME/CFS | 0.56 | 0.33 - 0.93 | **0.026** | 0.61 | 0.38 - 0.98 | **0.043** | 2.08 | 1.52 - 2.83 | **<0.001** | 0.51 | 0.28 - 0.93 | **0.028** |
| sr-IBS | 0.69 | 0.38 - 1.28 | 0.245 | 0.74 | 0.42 - 1.31 | 0.301 | 0.76 | 0.52 - 1.09 | 0.139 | 0.63 | 0.30 - 1.28 | 0.203 |
| Site 2 (Florida) | 1.99 | 0.80 - 4.97 | 0.141 | 1.63 | 0.70 - 3.82 | 0.262 | 2.09 | 1.21 - 3.62 | **0.009** | 1.79 | 0.61 - 5.22 | 0.288 |
| Site 3 (Nevada) | 1.41 | 0.65 - 3.06 | 0.382 | 1.14 | 0.56 - 2.35 | 0.715 | 1.38 | 0.87 - 2.20 | 0.172 | 1.82 | 0.74 - 4.50 | 0.195 |
| Site 4 (New York) | 1.24 | 0.60 - 2.57 | 0.554 | 1.21 | 0.61 - 2.37 | 0.586 | 0.86 | 0.56 - 1.33 | 0.510 | 0.95 | 0.41 - 2.23 | 0.915 |
| Site 5 (Utah) | 1.12 | 0.57 - 2.20 | 0.742 | 1.09 | 0.58 - 2.04 | 0.792 | 1.20 | 0.80 - 1.80 | 0.375 | 1.22 | 0.55 - 2.69 | 0.627 |
| Sex | 0.81 | 0.46 - 1.43 | 0.463 | 0.70 | 0.41 - 1.20 | 0.194 | 0.94 | 0.67 - 1.32 | 0.723 | 1.04 | 0.53 - 2.03 | 0.910 |
| BMI | 0.79 | 0.63 - 1.00 | 0.052 | 0.80 | 0.64 - 0.99 | **0.043** | 0.81 | 0.71 - 0.94 | **0.004** | 0.82 | 0.62 - 1.07 | 0.148 |
| Hispanic | 0.48 | 0.16 - 1.46 | 0.195 | 0.86 | 0.30 - 2.43 | 0.776 | 1.12 | 0.57 - 2.19 | 0.745 | 0.30 | 0.08 - 1.13 | 0.076 |
| Non-White & Non-Hispanic | 1.80 | 0.66 - 4.91 | 0.256 | 1.91 | 0.75 - 4.87 | 0.178 | 1.42 | 0.78 - 2.59 | 0.258 | 2.72 | 0.84 - 8.84 | 0.098 |
| Age | 0.99 | 0.78 - 1.27 | 0.951 | 0.98 | 0.78 - 1.23 | 0.864 | 1.10 | 0.95 - 1.27 | 0.200 | 1.17 | 0.88 - 1.55 | 0.289 |
| Antibiotics | 0.70 | 0.32 - 1.55 | 0.379 | 0.44 | 0.21 - 0.91 | **0.029** | 0.50 | 0.31 - 0.81 | **0.006** | 0.35 | 0.14 - 0.90 | **0.030** |
| Probiotics | 1.03 | 0.59 - 1.80 | 0.912 | 1.42 | 0.84 - 2.38 | 0.189 | 1.11 | 0.79 - 1.55 | 0.553 | 1.00 | 0.52 - 1.92 | 0.993 |
| Prebiotics | 0.67 | 0.25 - 1.74 | 0.408 | 0.73 | 0.30 - 1.79 | 0.494 | 0.72 | 0.40 - 1.28 | 0.259 | 0.61 | 0.20 - 1.88 | 0.391 |

Results for each outcome variable were estimated using generalized linear regression with a Gamma distribution with log link and adjusted for covariates.
Fold Change, 95% Confidence Intervals (CI), and p-values (*p*) are shown for each model.
Significant predictors (*p*<0.05) are indicated in bold.

The findings indicate that, despite having higher quantities of total bacterial 16S in feces, ME/CFS patients had significantly lower quantities of important BPB such as the genera *Roseburia* and *Eubacterium* and the species *F. prausnitzii*.

### Fecal bacterial functional metagenomic pathways differ between ME/CFS and healthy controls

Functional metagenomic analysis was carried out using GOmixer, a human gut microbiome-specific metabolic pathway analysis tool [40]. Comparison between ME/CFS subjects and healthy controls revealed nine global metabolic processes that differed between the gut microbiome of ME/CFS subjects and controls (**Figure 3A** and **Supplementary Table 3A**). Two processes, butyrate and sulfate metabolism, were deficient in ME/CFS. Seven processes, CO_2_ metabolism, monosaccharide degradation, acetogenesis, ethanol production, cross-feeding intermediate metabolism, sugar acid degradation and mucus degradation, were enriched in ME/CFS. At the level of gut metabolic modules, whereas six modules were deficient, fifteen modules were enriched (**Supplementary Figure 3** and **Supplementary Table 3B**). The specific module for *butyrate production via transferase* (MF0116) was deficient in ME/CFS, consistent with deficiencies in BPB we observed based on differential abundance and qPCR analyses. Other modules that were deficient in the ME/CFS microbiome included *superoxide reductase* (MF0132) and *sorbitol degradation* (MF0073). Modules that were enriched in ME/CFS included *lactate production* (MF0119), *pyruvate dehydrogenase complex* (MF0083), *ribose degradation* (MF0060), and *menaquinone production* (MF0133). Analysis of GOmixer gut-brain modules also revealed deficient metagenomic content for *Butyrate synthesis II* (MGB053), along with *ClpB (ATP-dependent chaperone protein)* (MGB029), and *S-Adenosylmethionine (SAM) synthesis* (MGB036) (**Supplementary Table 3C**). Enriched gut-brain modules in ME/CFS included *Menaquinone synthesis (vitamin K2) I* (MGB040), *GABA synthesis III* (MGB022), and *Isovaleric acid synthesis I (KADH pathway)* (MGB034).

**Figure 3:**
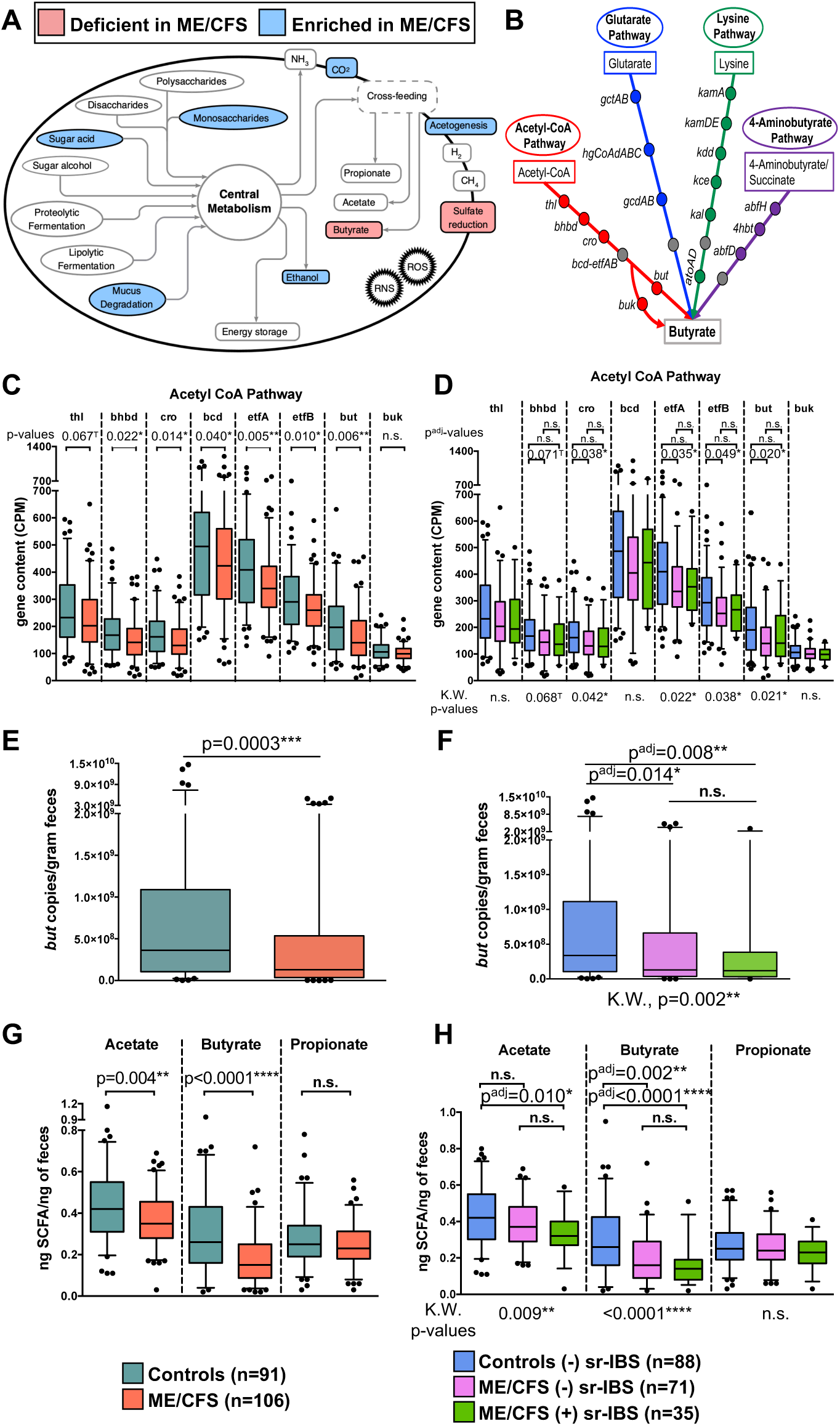
Functional metagenomic and metabolomic analyses. Gut bacterial metabolic processes that differ (deficient or enriched) in ME/CFS compared with healthy controls (**A**). Schematic showing the 4 gut bacterial pathways of butyrate production and the bacterial genes involved in the production of butyrate (**B**). Note: gray circles indicate that the bcd-etfAB complex is shared among all 4 pathways. Box-and-whiskers plots showing the distribution of gene counts (CPM) for genes in the acetyl-CoA pathway of butyrate production (**C, D**) between ME/CFS and healthy controls (**C**) and among stratified groups (**D**). Box-and-whiskers plots showing the distribution of fecal *but* gene copies per gram of feces determined by qPCR between ME/CFS and healthy controls (**E**) and among stratified groups (**F**). Box-and-whiskers plots showing the distribution of fecal SCFAs measured by GC-MS between ME/CFS and healthy controls (**G**) and among stratified groups (**H**). Statistical significance was determined based on two-tailed p-values from the Mann-Whitney U test for each gene (**C, E, G**). For stratified analyses significance was first determined based on the Kruskal-Wallis test for each gene (K.W., results shown below each figure in **D, F, H**). If significant (p < 0.05) based on K.W., then between group significance was determined based on the Mann-Whitney U test with multiple testing (Bonferroni) correction (p^adj^-value). n.s. = not significant; * = p or p^adj^ < 0.05; ** = p or p^adj^ < 0.01; *** = p or p^adj^ < 0.001; **** = p or p^adj^ < 0.0001; T = trend (p or p^adj^ < 0.1).

As differential abundance analysis of taxa, qPCR, and functional metagenomic analysis implicated deficiencies in BPB and functional capacity of the gut microbiome to produce butyrate in ME/CFS subjects relative to healthy controls, we investigated the metagenomic gene content for genes along the four pathways of gut bacterial butyrate production: the acetyl-CoA pathway, the glutarate pathway, the lysine pathway, and the 4-aminobutyrate pathway (**Figure 3B**). We aligned bacterial reads to a curated database of genes involved in butyrate production along the four pathways [41]. Comparing the metagenomic content (Counts per Million, CPM) of each gene along the acetyl-CoA pathway showed deficient gene content for nearly every gene along this pathway in ME/CFS relative to healthy controls (Mann-Whitney U test, p < 0.05 for all genes except *thl* and *buk*) (**Figure 3C**). Quantitatively, the majority of bacteria in the intestine encode and utilize *but* (butyryl-CoA:acetate CoA transferase) rather than *buk* (butyrate kinase) as the terminal gene in the acetyl-CoA pathway to produce butyrate. In fact, most bacteria of the *Eubacterium*, *Roseburia*, and *Faecalibacterium* genera (those we found to be deficient in ME/CFS) only encode the *but* gene [41]. While nearly all genes of the acetyl-CoA pathway differed between ME/CFS and healthy controls, only terminal genes of the glutarate pathway (*gcdA* and *gcdB*), and both early and terminal genes of the lysine pathway (*kamA*, *kamD*, *kamE*, *atoA*, *atoD*) were deficient in ME/CFS compared to healthy controls, and no genes differed in the 4-aminobutyrate pathway in univariate analyses **(Supplementary Figure 4A-C**). Even in stratified analyses by sr-IBS status, most genes along the acetyl-CoA pathway remained significantly depleted in the microbiome of ME/CFS subjects without sr-IBS compared with healthy controls without sr-IBS (Mann-Whitney U test, p^adj^ = 0.071, 0.038, 0.035, 0.049, 0.020 for *bhbd*, *cro*, *etfA*, *etfB*, and *but*, respectively) (**Figure 3D**). In contrast, no genes along the glutarate, lysine, or 4- aminobutyrate pathways differed significantly among groups in stratified analyses (**Supplementary Figure 4D-F**).

We further evaluated metagenomic gene content for genes along butyrate pathways using generalized linear regression with a Gamma distribution with log link and adjusting for covariates. Even after adjusting for covariates, ME/CFS was a significant predictor of most genes along the acetyl-CoA pathway (all except *thl* and *buk*) (**Table 4**). Specifically, quantities of *bhbd*, *cro*, *bcd*, *etfA*, *etfB* and *but* were 0.81-fold (95% CI: 0.68-0.96, p = 0.014), 0.80-fold (95% CI: 0.68-0.95, p = 0.008), 0.85-fold (95% CI: 0.73-1.00, p = 0.043), 0.84-fold (95% CI: 0.74-0.95, p = 0.005), 0.85-fold (95% CI: 0.75-0.96, p = 0.007) and 0.75-fold (95% CI: 0.62-0.91, p = 0.003) lower in ME/CFS than in controls, respectively. In addition to ME/CFS status, BMI was associated with only *but* gene content. In contrast, only a few genes in the glutarate pathway were associated with ME/CFS status (*gctA*, *hgCoAdC* and *gcdA*) in regression analyses (**Supplementary Table 4A**). Only the terminal genes, *atoA* and *atoD*, were associated with ME/CFS status in the lysine pathway. Interestingly, most genes along the lysine pathway were independently associated with race/ethnicity (**Supplementary Table 4B**). None of the genes in the 4-aminobutyrate pathway were significantly associated with ME/CFS status (**Supplementary Table 4C**).

**Table 4:** Generalized linear regression showing the relationship between ME/CFS status and metagenomic gene content in the acetyl-CoA pathway.

(Part-1/2)
| *Predictors* | Acetyl-CoA pathway |  |  |  |  |  |  |  |  |  |  |  |
| --- | --- | --- | --- | --- | --- | --- | --- | --- | --- | --- | --- | --- |
|  | thl |  |  | bhbd |  |  | cro |  |  | bcd |  |  |
|  | *Fold Change* | *95% CI* | *p* | *Fold Change* | *95% CI* | *p* | *Fold Change* | *95% CI* | *p* | *Fold Change* | *95% CI* | *p* |
| (Intercept) | 275.78 | 216 – 353 | **<0.001** | 203.9 | 160 – 260 | **<0.001** | 192.18 | 152 – 244 | **<0.001** | 523.23 | 420 – 655 | **<0.001** |
| ME/CFS | 0.86 | 0.72 – 1.03 | 0.087 | 0.81 | 0.68 – 0.96 | **0.014** | 0.80 | 0.68 – 0.95 | **0.008** | 0.85 | 0.73 – 1.00 | **0.043** |
| sr-IBS | 1.02 | 0.83 – 1.25 | 0.874 | 1.08 | 0.88 – 1.32 | 0.465 | 1.07 | 0.88 – 1.31 | 0.473 | 1.06 | 0.88 – 1.28 | 0.522 |
| Site 2 (Florida) | 1.04 | 0.77 – 1.41 | 0.806 | 1.03 | 0.76 – 1.39 | 0.868 | 1.01 | 0.75 – 1.35 | 0.970 | 0.97 | 0.74 – 1.28 | 0.839 |
| Site 3 (Nevada) | 1.06 | 0.83 – 1.36 | 0.634 | 0.99 | 0.78 – 1.27 | 0.964 | 1.01 | 0.79 – 1.28 | 0.951 | 1.00 | 0.80 – 1.25 | 0.987 |
| Site 4 (New York) | 1.01 | 0.79 – 1.28 | 0.962 | 0.97 | 0.76 – 1.23 | 0.795 | 0.96 | 0.76 – 1.22 | 0.753 | 0.98 | 0.78 – 1.21 | 0.819 |
| Site 5 (Utah) | 1.02 | 0.81 – 1.27 | 0.877 | 0.98 | 0.78 – 1.21 | 0.828 | 0.98 | 0.79 – 1.21 | 0.837 | 0.99 | 0.81 – 1.21 | 0.915 |
| Sex | 0.92 | 0.76 – 1.10 | 0.356 | 0.87 | 0.72 – 1.05 | 0.149 | 0.89 | 0.75 – 1.07 | 0.227 | 0.96 | 0.81 – 1.14 | 0.670 |
| BMI | 0.94 | 0.87 – 1.02 | 0.110 | 0.94 | 0.87 – 1.01 | 0.103 | 0.96 | 0.89 – 1.03 | 0.221 | 0.97 | 0.91 – 1.04 | 0.426 |
| Hispanic | 0.98 | 0.68 – 1.46 | 0.929 | 0.95 | 0.66 – 1.41 | 0.802 | 0.96 | 0.67 – 1.41 | 0.821 | 1.12 | 0.80 – 1.61 | 0.499 |
| Non-White & Non-Hispanic | 0.96 | 0.69 – 1.36 | 0.796 | 1.00 | 0.73 – 1.41 | 0.993 | 0.97 | 0.71 – 1.37 | 0.874 | 0.95 | 0.71 – 1.30 | 0.736 |
| Age | 1.01 | 0.93 – 1.09 | 0.790 | 1.00 | 0.93 – 1.09 | 0.926 | 1.00 | 0.93 – 1.08 | 0.907 | 1.01 | 0.94 – 1.09 | 0.695 |
| Antibiotics | 0.90 | 0.69 – 1.18 | 0.410 | 0.95 | 0.74 – 1.25 | 0.703 | 0.92 | 0.71 – 1.19 | 0.499 | 0.86 | 0.68 – 1.11 | 0.224 |
| Probiotics | 0.99 | 0.82 – 1.19 | 0.876 | 1.00 | 0.83 – 1.21 | 0.998 | 1.00 | 0.84 – 1.20 | 0.957 | 1.03 | 0.87 – 1.22 | 0.709 |
| Prebiotics | 1.08 | 0.80 – 1.50 | 0.622 | 1.07 | 0.79 – 1.48 | 0.654 | 1.06 | 0.79 – 1.45 | 0.712 | 1.04 | 0.79 – 1.40 | 0.795 |

(Part-2/2)
| *Predictors* | Acetyl-CoA pathway |  |  |  |  |  |  |  |  |  |  |  |
| --- | --- | --- | --- | --- | --- | --- | --- | --- | --- | --- | --- | --- |
|  | etfA |  |  | etfB |  |  | but |  |  | buk |  |  |
|  | *Fold Change* | *95% CI* | *p* | *Fold Change* | *95% CI* | *p* | *Fold Change* | *95% CI* | *p* | *Fold Change* | *95% CI* | *p* |
| (Intercept) | 455.1 | 383 – 542 | **<0.001** | 330.2 | 278 – 392 | **<0.001** | 237.18 | 182 – 311 | **<0.001** | 114.07 | 98 – 132 | **<0.001** |
| ME/CFS | 0.84 | 0.74 – 0.95 | **0.005** | 0.85 | 0.75 – 0.96 | **0.007** | 0.75 | 0.62 – 0.91 | **0.003** | 0.93 | 0.84 – 1.04 | 0.180 |
| sr-IBS | 1.03 | 0.89 – 1.19 | 0.731 | 1.03 | 0.89 – 1.19 | 0.675 | 1.08 | 0.87 – 1.36 | 0.493 | 0.94 | 0.83 – 1.07 | 0.345 |
| Site 2 (Florida) | 0.97 | 0.78 – 1.20 | 0.759 | 0.96 | 0.78 – 1.19 | 0.713 | 0.97 | 0.70 – 1.36 | 0.869 | 0.91 | 0.76 – 1.10 | 0.339 |
| Site 3 (Nevada) | 0.99 | 0.83 – 1.18 | 0.879 | 0.99 | 0.83 – 1.18 | 0.903 | 0.95 | 0.72 – 1.25 | 0.720 | 1.08 | 0.93 – 1.26 | 0.339 |
| Site 4 (New York) | 0.91 | 0.77 – 1.08 | 0.274 | 0.92 | 0.77 – 1.09 | 0.311 | 0.94 | 0.72 – 1.22 | 0.644 | 0.87 | 0.75 – 1.01 | 0.061 |
| Site 5 (Utah) | 0.97 | 0.83 – 1.14 | 0.713 | 0.98 | 0.84 – 1.14 | 0.776 | 0.96 | 0.75 – 1.23 | 0.773 | 0.98 | 0.86 – 1.12 | 0.782 |
| Sex | 0.92 | 0.81 – 1.05 | 0.244 | 0.94 | 0.82 – 1.07 | 0.332 | 0.86 | 0.70 – 1.06 | 0.165 | 0.99 | 0.88 – 1.10 | 0.804 |
| BMI | 0.99 | 0.94 – 1.05 | 0.810 | 1.00 | 0.95 – 1.05 | 0.976 | 0.92 | 0.84 – 1.00 | 0.045 | 1.02 | 0.97 – 1.07 | 0.389 |
| Hispanic | 1.08 | 0.83 – 1.42 | 0.556 | 1.09 | 0.84 – 1.43 | 0.503 | 0.89 | 0.60 – 1.37 | 0.563 | 1.05 | 0.84 – 1.33 | 0.660 |
| Non-White & Non-Hispanic | 0.91 | 0.72 – 1.16 | 0.411 | 0.91 | 0.73 – 1.16 | 0.430 | 1.09 | 0.77 – 1.60 | 0.637 | 0.92 | 0.76 – 1.14 | 0.437 |
| Age | 1.02 | 0.96 – 1.08 | 0.582 | 1.02 | 0.96 – 1.08 | 0.535 | 0.99 | 0.90 – 1.08 | 0.760 | 1.02 | 0.97 – 1.07 | 0.414 |
| Antibiotics | 0.94 | 0.78 – 1.14 | 0.528 | 0.93 | 0.78 – 1.13 | 0.453 | 0.99 | 0.75 – 1.34 | 0.966 | 0.97 | 0.83 – 1.15 | 0.754 |
| Probiotics | 1.02 | 0.89 – 1.16 | 0.790 | 1.03 | 0.90 – 1.17 | 0.669 | 1.03 | 0.84 – 1.26 | 0.763 | 0.98 | 0.87 – 1.09 | 0.665 |
| Prebiotics | 1.07 | 0.86 – 1.34 | 0.577 | 1.07 | 0.87 – 1.34 | 0.532 | 0.94 | 0.68 – 1.34 | 0.733 | 1.13 | 0.93 – 1.38 | 0.216 |

Results for each outcome variable were estimated using generalized linear regression with a Gamma distribution with log link and adjusted for covariates.
Fold Change, 95% Confidence Intervals (CI), and p-values (p) are shown for each model.
Significant predictors (p<0.05) are indicated in bold.

### Quantitation of the bacterial *but* gene in feces

*But*, as the dominant terminal gene in the bacterial acetyl-CoA pathway for butyrate production, can serve as an indicator gene for the overall butyrate-producing capacity in the human gut [42]. We quantitated the *but* gene in fecal samples from ME/CFS and control subjects by qPCR. The overall *but* copies/gram feces were lower in ME/CFS compared to control subjects (Mann-Whitney U test, p = 0.0003) (**Figure 3E**). Further, stratified analyses by sr-IBS status revealed lower *but* copies/gram feces in ME/CFS without sr-IBS (Mann-Whitney U test, p^adj^ = 0.014) and ME/CFS with sr-IBS (Mann-Whitney U test, p^adj^ = 0.008) compared to healthy controls without sr-IBS (**Figure 3F**). No differences were found comparing ME/CFS subjects with and without sr-IBS. These results suggest that deficient *but* gene quantity in the feces of ME/CFS patients is independent of sr-IBS status.

We also assessed the relative abundance of *but* gene copies to total bacterial 16S in feces based on qPCR. As with the total quantity of *but* gene/gram of feces, the *but* gene calculated relative abundance was lower in ME/CFS subjects compared to healthy controls in univariate analyses (Mann-Whitney U test, p = 0.0003) (**Supplementary Figure 5A**). In stratified analyses by sr-IBS status, *but* gene relative abundance was lower in ME/CFS subjects without sr-IBS compared with healthy controls without sr-IBS (Mann-Whitney U test, p^adj^ = 0.002). A trend toward lower *but* gene relative abundance was observed in ME/CFS subjects with sr-IBS compared with healthy controls without sr-IBS (Mann-Whitney U test, p^adj^ = 0.096) **(Supplementary Figure 5B**).

To further assess the relationships between quantitative measures of the *but* gene in feces (outcome variable) with ME/CFS status (main predictor) generalized linear regression models with a Gamma distribution with log link were assessed, adjusting for covariates (**Table 3**). Even after adjusting for covariates in the model, ME/CFS was associated with *but* gene quantities (FC=0.51, 95% CI: 0.28-0.93, p = 0.028). Of the covariates, only antibiotic usage in the prior 6-12 weeks showed an independent association with *but* gene quantity. Similarly, lower calculated relative abundance of the *but* gene was significantly associated with ME/CFS status (FC=0.60, 95% CI: 0.39-0.91, p = 0.017) in regression and only Hispanic race showed an independent association with *but* gene relative abundance in the model (**Supplementary Table 2**).

These results confirm quantitative deficiency of the most important terminal gene in the acetyl-CoA pathway of butyrate production in the feces of ME/CFS subjects compared to healthy controls.

### Assessment of SCFAs in feces

Given the strong evidence indicating reduced abundance and quantity of BPB and reduced metagenomic capacity for producing butyrate, we measured SCFAs in all fecal samples from ME/CFS and healthy control subjects using gas chromatography-mass spectrometry. The fecal concentration of both acetate (Mann-Whitney U test, p = 0.004) and butyrate (Mann-Whitney U test, p < 0.0001) were lower in ME/CFS compared to healthy controls, while propionate was unchanged, in univariate analyses (**Figure 3G**). In stratified analyses by sr-IBS status, acetate was lower in ME/CFS subjects with sr-IBS compared to healthy controls without sr-IBS (Mann-Whitney U test, p^adj^ = 0.010), but did not differ in ME/CFS subjects without sr-IBS. In contrast, butyrate was lower in both ME/CFS subjects without sr-IBS (Mann-Whitney U test, p^adj^ < 0.0001) and with sr-IBS (Mann-Whitney U test, p^adj^ = 0.002) compared to healthy controls without sr-IBS (**Figure 3H**).

To further assess the relationships between quantitative measures of SCFAs in feces (outcome variables) with ME/CFS status (main predictor), generalized linear regression models with Gamma distribution with log link were fitted, adjusting for covariates (**Table 5**). As with all our regression analyses, we evaluated the interaction term between ME/CFS and sr-IBS. Unlike our prior regressions, the ME/CFS*sr-IBS interaction term was significant. In order to better understand the nature of the relationships among SCFA levels and ME/CFS and sr-IBS, we evaluated stratified regression models, adjusted for covariates (bottom 3 rows in **Table 5**, and **Supplementary Tables 5A, 5B, 5C**). Regression Model 2, comparing levels of SCFAs between ME/CFS subjects without sr-IBS and healthy controls without sr-IBS, revealed decreased quantities of both acetate (FC=0.87, 95% CI: 0.77-0.98, p=0.022) and butyrate (FC=0.60, 95% CI: 0.48-0.75, p<0.001), but not propionate (FC=0.90, 95% CI: 0.78-1.04, p=0.149) in ME/CFS. Regression Model 3, comparing ME/CFS subjects with sr-IBS and healthy controls without sr-IBS, revealed decreased quantities of acetate (FC=0.83, 95% CI: 0.71-0.96, p=0.017), butyrate (FC=0.56, 95% CI: 0.42-0.73, p<0.001) and propionate (FC=0.83, 95% CI: 0.70-1.00, p=0.047) in ME/CFS subjects with IBS. Regression Model 4, comparing ME/CFS subjects with sr-IBS and ME/CFS subjects without sr-IBS, did not identify differences in any of the SCFAs between these groups. These results indicate that both deficient acetate and butyrate are associated with ME/CFS with or without comorbid sr-IBS, while deficient propionate is only found in ME/CFS subjects with sr-IBS. Further, there is a larger effect size in terms of the estimated fold change for fecal butyrate concentrations compared with either acetate or propionate.

**Table 5:**
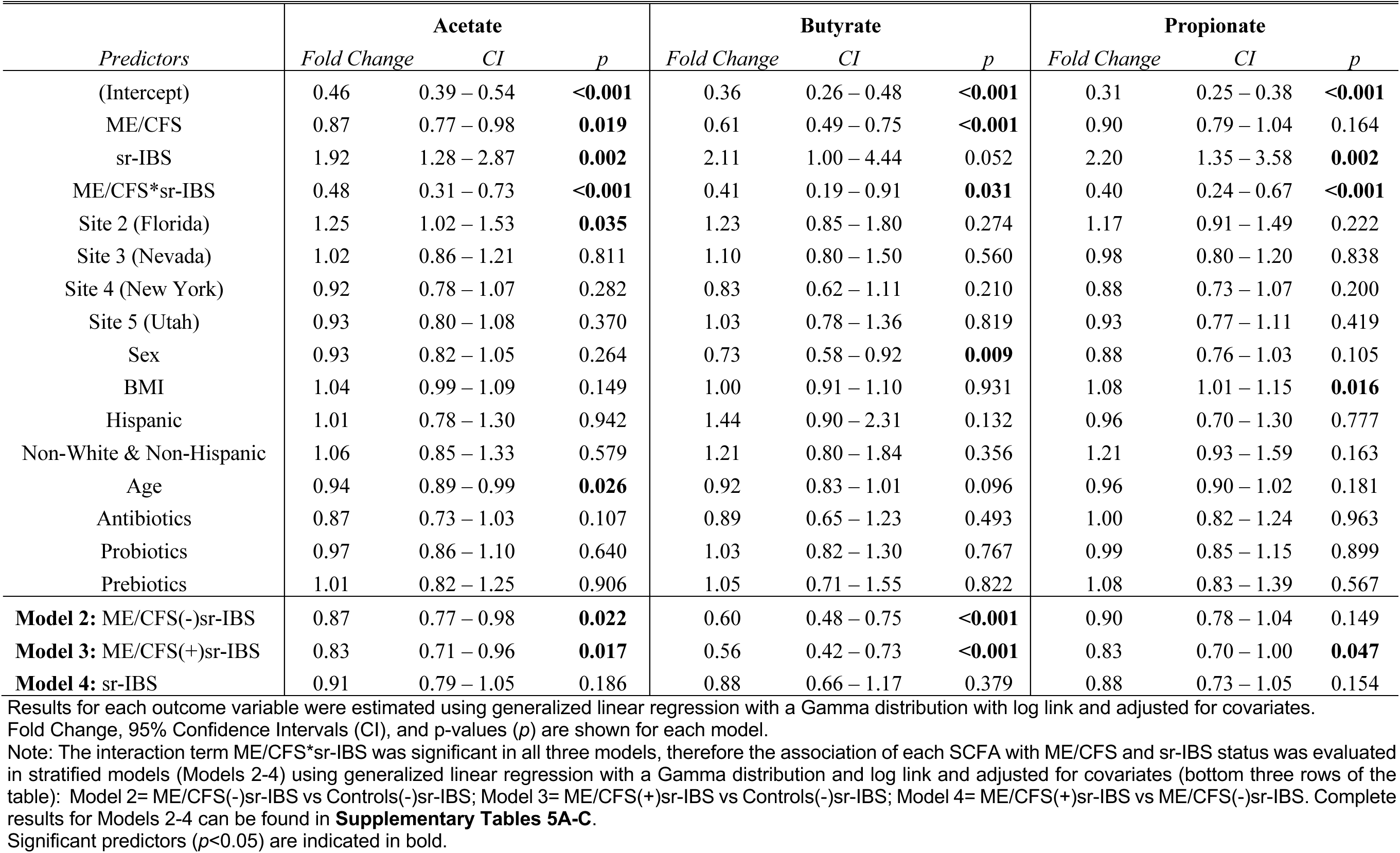
Generalized linear regression showing the relationship between ME/CFS status and SCFAs.

| *Predictors* | **Acetate** |  |  | **Butyrate** |  |  | **Propionate** |  |  |
| --- | --- | --- | --- | --- | --- | --- | --- | --- | --- |
|  | *Fold Change* | *CI* | *p* | *Fold Change* | *CI* | *p* | *Fold Change* | *CI* | *p* |
| (Intercept) | 0.46 | 0.39 – 0.54 | **<0.001** | 0.36 | 0.26 – 0.48 | **<0.001** | 0.31 | 0.25 – 0.38 | **<0.001** |
| ME/CFS | 0.87 | 0.77 – 0.98 | **0.019** | 0.61 | 0.49 – 0.75 | **<0.001** | 0.90 | 0.79 – 1.04 | 0.164 |
| sr-IBS | 1.92 | 1.28 – 2.87 | **0.002** | 2.11 | 1.00 – 4.44 | 0.052 | 2.20 | 1.35 – 3.58 | **0.002** |
| ME/CFS*sr-IBS | 0.48 | 0.31 – 0.73 | **<0.001** | 0.41 | 0.19 – 0.91 | **0.031** | 0.40 | 0.24 – 0.67 | **<0.001** |
| Site 2 (Florida) | 1.25 | 1.02 – 1.53 | **0.035** | 1.23 | 0.85 – 1.80 | 0.274 | 1.17 | 0.91 – 1.49 | 0.222 |
| Site 3 (Nevada) | 1.02 | 0.86 – 1.21 | 0.811 | 1.10 | 0.80 – 1.50 | 0.560 | 0.98 | 0.80 – 1.20 | 0.838 |
| Site 4 (New York) | 0.92 | 0.78 – 1.07 | 0.282 | 0.83 | 0.62 – 1.11 | 0.210 | 0.88 | 0.73 – 1.07 | 0.200 |
| Site 5 (Utah) | 0.93 | 0.80 – 1.08 | 0.370 | 1.03 | 0.78 – 1.36 | 0.819 | 0.93 | 0.77 – 1.11 | 0.419 |
| Sex | 0.93 | 0.82 – 1.05 | 0.264 | 0.73 | 0.58 – 0.92 | **0.009** | 0.88 | 0.76 – 1.03 | 0.105 |
| BMI | 1.04 | 0.99 – 1.09 | 0.149 | 1.00 | 0.91 – 1.10 | 0.931 | 1.08 | 1.01 – 1.15 | **0.016** |
| Hispanic | 1.01 | 0.78 – 1.30 | 0.942 | 1.44 | 0.90 – 2.31 | 0.132 | 0.96 | 0.70 – 1.30 | 0.777 |
| Non-White & Non-Hispanic | 1.06 | 0.85 – 1.33 | 0.579 | 1.21 | 0.80 – 1.84 | 0.356 | 1.21 | 0.93 – 1.59 | 0.163 |
| Age | 0.94 | 0.89 – 0.99 | **0.026** | 0.92 | 0.83 – 1.01 | 0.096 | 0.96 | 0.90 – 1.02 | 0.181 |
| Antibiotics | 0.87 | 0.73 – 1.03 | 0.107 | 0.89 | 0.65 – 1.23 | 0.493 | 1.00 | 0.82 – 1.24 | 0.963 |
| Probiotics | 0.97 | 0.86 – 1.10 | 0.640 | 1.03 | 0.82 – 1.30 | 0.767 | 0.99 | 0.85 – 1.15 | 0.899 |
| Prebiotics | 1.01 | 0.82 – 1.25 | 0.906 | 1.05 | 0.71 – 1.55 | 0.822 | 1.08 | 0.83 – 1.39 | 0.567 |
| **Model 2:** ME/CFS(-)sr-IBS | 0.87 | 0.77 – 0.98 | **0.022** | 0.60 | 0.48 – 0.75 | **<0.001** | 0.90 | 0.78 – 1.04 | 0.149 |
| **Model 3:** ME/CFS(+)sr-IBS | 0.83 | 0.71 – 0.96 | **0.017** | 0.56 | 0.42 – 0.73 | **<0.001** | 0.83 | 0.70 – 1.00 | **0.047** |
| **Model 4:** sr-IBS | 0.91 | 0.79 – 1.05 | 0.186 | 0.88 | 0.66 – 1.17 | 0.379 | 0.88 | 0.73 – 1.05 | 0.154 |

Results for each outcome variable were estimated using generalized linear regression with a Gamma distribution with log link and adjusted for covariates. Fold Change, 95% Confidence Intervals (CI), and p-values (*p*) are shown for each model.
Note: The interaction term ME/CFS\*sr-IBS was significant in all three models, therefore the association of each SCFA with ME/CFS and sr-IBS status was evaluated in stratified models (Models 2-4) using generalized linear regression with a Gamma distribution and log link and adjusted for covariates (bottom three rows of the table): Model 2= ME/CFS(-)sr-IBS vs Controls(-)sr-IBS; Model 3= ME/CFS(+)sr-IBS vs Controls(-)sr-IBS; Model 4= ME/CFS(+)sr-IBS vs ME/CFS(-)sr-IBS. Complete results for Models 2-4 can be found in **Supplementary Tables 5A-C**.
Significant predictors ( $p < 0.05$ ) are indicated in bold.

### Relationships among measured variables and fatigue scores

Relative abundance, quantitative, and functional analyses of microbiota were consistent and highly correlated even after FDR adjustment for Spearman’s correlations (**Figure 4A**). Most measured factors showed positive correlations despite a wide range of laboratory methods employed (metagenomics analysis of taxa relative abundance and alpha diversity, metagenomics analysis of genes in the acetyl-CoA pathway, qPCR analysis of bacterial taxa and the *but* gene, and GC-MS measures of stool SCFAs including butyrate). Some of the stronger positive correlations such as those observed between the relative abundance of *F. prausnitzii* determined by shotgun metagenomics and the quantity and relative abundance of *F. prausnitzii* determined by qPCR, speak to the reproducibility of our findings using different methods. Other positive correlations, such as those among alpha diversity, gene counts along the acetyl-CoA pathway (especially *but*) and the relative abundance or quantity of BPB may be more reflective of biological links among diversity and function of bacteria. Positive correlations among measured variables were generally reflective of measured variables that tended to be deficient in ME/CFS or ME/CFS stratified by sr-IBS, which was the case for the majority of measured variables evaluated here. Inverse correlations were generally restricted to relative abundance of bacterial taxa that were found to be enriched in ME/CFS (*Lachnoclostridium*) or ME/CFS with sr-IBS (*C. bolteae*, *F. plautii*, *Flavonifractor*).

**Figure 4:**
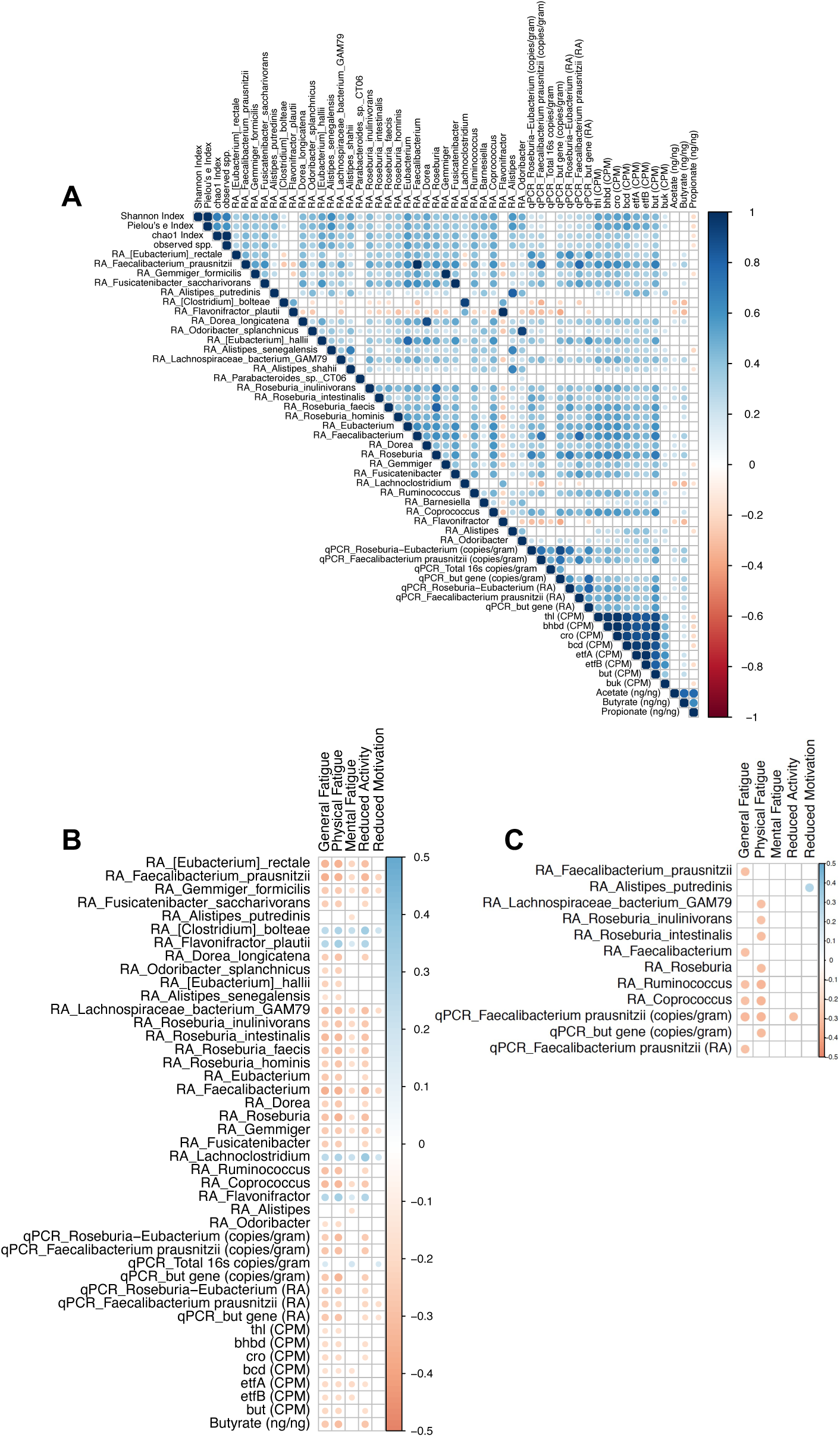
Spearman’s correlation analysis among measured variables and fatigue scores. Spearman’s correlation assessing relationships among all measured variables in both cases and controls (n=197) (**A**), and between measured variables and fatigue dimension scores in both cases and controls (n=197) (**B**), and only within ME/CFS cases (n=106) (**C**). Note: no significant correlations between measured variables and fatigue scores were observed within healthy controls (n=91), only within ME/CFS cases without sr-IBS (n=71), or only within ME/CFS cases with sr-IBS (n=35). Each colored circle in each correlogram represents a significant correlation (nonsignificant correlations are white). The color bars show the scale for the Spearman’s rank correlation coefficient. Both the size and color of the circles indicate the strength of the correlation. FDR was controlled with the Benjamini-Hochberg procedure, with correlations with adjusted p-values < 0.05 shown. In figures B and C, all measured variables (shown in **A**) were compared against all five fatigue dimensions, and only significant correlations are shown (all other comparisons were not significant after adjustment).

We also evaluated the relationship among our measured variables and fatigue scores based on the five dimensions of the Multidimensional Fatigue Inventory (MFI) on the whole dataset (all cases + all controls, n=197) (**Figure 4B)**. In this case, the majority of significant correlations among measured variables and fatigue scores after FDR correction were inverse, as measured variables that tended to be deficient in ME/CFS were associated with higher fatigue scores (more severe fatigue), especially for general and physical fatigue and reduced activity. The relative abundance and qPCR quantitation of bacterial species and the *but* gene, metagenomic content of genes along the acetyl-CoA butyrate pathway, and levels of butyrate were inversely correlated with general fatigue, physical fatigue and reduced activity.

As correlations in the whole dataset may reflect reduced levels of measured variables and higher fatigue scores in individuals with ME/CFS vs. healthy controls (**Supplementary Table 6**), we also evaluated associations between measured variables and fatigue scores in ME/CFS subjects alone (n=106) (**Figure 4C**). Although fewer correlations remained significant in the cases after FDR adjustment, various bacterial taxa including *F. prausnitzii*, the genus *Faecalibacterium*, *Roseburia inulinivorans*, *Roseburia intestinalis*, the genus *Roseburia*, and the genus *Coprococcus* correlated inversely with either general fatigue, or physical fatigue, or both. The qPCR quantity for the *but* gene correlated inversely with general fatigue and the qPCR quantity of *F. prausnitzii* in stool correlated inversely with general fatigue, physical fatigue and reduced activity (**Figure 4C**). Thus, the lower the abundance or quantity of these BPB, the more severe the fatigue dimension scores in ME/CFS.

We also determined whether similar relationships existed among our measured variables and fatigue scores in healthy control subjects alone (n=91). In contrast to ME/CFS subjects, no significant correlations were found in healthy controls after FDR adjustment (as such, no correlogram is shown).

As sr-IBS could further influence these relationships, we evaluated the relationships among measured variables and fatigue scores only in ME/CFS patients without sr-IBS (n=71) and only in ME/CFS patients with sr-IBS (n=35). No significant relationships were found for either stratified group after FDR adjustment (as such, no correlograms are shown).

Overall, these findings suggest that deficiencies in BPB and their metabolic pathways are correlated with the severity of fatigue symptoms, but these associations are only found in ME/CFS, not healthy controls.

## Discussion

Our analyses of the fecal microbiome in ME/CFS subjects and healthy controls matched for age, sex, geography, and socioeconomic status indicated significant differences in composition, function and metabolism. In a systematic review of studies on gut dysbiosis in ME/CFS, Du Preez et al. concluded that intrinsic and extrinsic factors that can influence microbiome composition should be better controlled for in case-control studies [24]. One such factor that may be important in the context of ME/CFS and the microbiome is IBS co-morbidity, which occurs at high prevalence in ME/CFS compared with healthy controls [31–33], and is a condition that is independently associated with microbiome disturbances [35, 43, 44]. In our study, more ME/CFS patients reported having an IBS diagnosis (35/106; 33%) than healthy controls (3/91; 3.3%). Both our current and prior fecal shotgun metagenomic study support the notion that IBS co-morbidity must be carefully considered in future ME/CFS microbiome studies. In fact, differences in gut microbiome alpha diversity between ME/CFS subjects and healthy controls appears to be largely driven by sr-IBS co-morbidity in ME/CFS, and individuals with ME/CFS and sr-IBS had a range of distinct bacterial species with differential relative abundance compared to healthy controls (i.e., *A. putredinis*, *C. bolteae*, *F. plautii*, *D. longicatena*) that were not found when comparing ME/CFS without sr-IBS and healthy controls. Thus, at least some microbiome differences in ME/CFS are confounded by IBS co-morbidity. In contrast, microbiota beta diversity differed significantly between ME/CFS and healthy controls; a difference that was independent of sr-IBS status.

### Species and taxa linked to ME/CFS: Relative and Absolute Differences

GAMLSS-BEZI models, adjusted for important covariates (including sr-IBS), identified differentially abundant fecal bacterial taxa between ME/CFS subjects and healthy controls. These analyses identified two species (*E. rectale* and *F. prausnitzii*) and six genera (*Eubacterium*, *Faecalibacterium*, *Dorea*, *Roseburia*, *Gemmiger*, and *Lachnoclostridium*) that differed in relative abundance in ME/CFS compared to healthy control subjects. Both the species *E. rectale* and *F. prausnitzii* and the genera *Eubacterium*, *Faecalibacterium* and *Roseburia* are prominent BPB in the human GI tract [41]. All had lower relative abundance in feces from ME/CFS subjects compared to healthy controls, independent of sr-IBS status. To test the validity of this finding, we employed qPCR to quantitatively evaluate levels of *Roseburia*-*Eubacterium* and *F. prausnitzii*. Both the fecal load of these BPB as well as their relative abundances were deficient in fecal samples from ME/CFS patients.

An additional, unexpected and novel finding from our qPCR analyses was that ME/CFS patients have higher total bacterial 16S rRNA genes/gram of feces than healthy controls. Total fecal bacterial load is infrequently evaluated in microbiome studies and little is still known about factors that may impact total bacterial load. However, it is well documented that antibiotics can have a dramatic impact on bacterial load [45, 46]. In this study, we controlled for antibiotics, both in our study design and statistical analyses. Thus, it is unlikely that antibiotics can explain the differences in bacterial load.

Dietary factors that may impact bacterial load include low fermentable oligo-, di-, mono-saccharides and polyols (FODMAPs) [47]. However, we cannot address this possibility because detailed dietary information was not collected from subjects. While it is possible that subject diets differ between ME/CFS subjects and controls, we note that more ME/CFS subjects reported taking prebiotic fiber supplements (11/106; 10.4%) than healthy controls (2/91; 2.2%). Prebiotic fibers typically stimulate the growth and activity of BPB. The observation that ME/CFS subjects had lower levels of BPB and butyrate and higher total bacterial load even with adjustment of regression models for prebiotic fiber use, suggests that some factor, either associated with the pathobiology or symptoms of ME/CFS, rather than prebiotic fiber intake is selectively influencing these microbiome changes.

Other gastrointestinal disturbances such as severe acute malnutrition with acute diarrhea and inflammatory bowel disease can also alter total bacterial load in feces or intestinal mucosa, respectively [48, 49]. However, subjects in this study were not acutely malnourished and only two cases (2/106) reported a formal diagnosis of IBD; no controls had a diagnosis of IBD. Small intestinal bacterial overgrowth is frequently associated with functional gastrointestinal disorders such as IBS [50]. However, we are not aware of any studies that have shown that fecal bacterial load is increased in IBS, and our findings suggest that increased fecal bacterial load is associated with ME/CFS, independent of sr-IBS. Further, despite the significant association of fecal bacterial load and ME/CFS, it remains unclear whether higher fecal bacterial load is representative of bacterial overgrowth in the intestine or a higher rate of bacterial washout or loss of adherent mucosa-associated bacteria.

### Specific butyrate-production deficiency in ME/CFS

Our functional metagenomic analysis found that the overall bacterial capacity for butyrate production is deficient in ME/CFS. To delve deeper into the specific pathways of butyrate production that are deficient, we examined the bacterial gene content of the four bacterial butyrate production pathways. Only the gene content for the acetyl-CoA pathway was deficient in ME/CFS. The acetyl-CoA pathway is the dominant pathway of butyrate production in the human gut. Whereas this pathway for butyrate production is fueled by carbohydrates, the other three pathways (glutarate, lysine and 4-aminobutyrate pathways) are fueled by proteins [41]. Counts per million of genes in the acetyl-CoA pathway were substantially higher than for genes in the other three pathways.

The majority of bacteria that utilize the acetyl-CoA pathway typically encode either the *but* gene or the *buk* gene to complete the terminal reaction of butyrate production. The *but* gene is dominant in the human gut [41]. We found that whereas the *but* gene was deficient in ME/CFS, the *buk* gene was not. Several of the bacteria that we found at lower relative abundance and quantity in ME/CFS, including *F. prausnitzii*, *E. rectale*, *Roseburia* spp. and *Eubacterium* spp. are known to encode the genes for the acetyl-CoA pathway of butyrate [41]. Our functional metagenomic analyses showed that ME/CFS subjects are specifically deficient in BPB that rely on the terminal *but* gene to produce butyrate. qPCR and metabolomic analyses corroborated our functional metagenomic findings by confirming that the quantity of the bacterially encoded *but* genes and levels of butyrate are lower in feces of ME/CFS patients than in healthy controls.

In addition to butyrate, ME/CFS subjects have reduced quantities of fecal acetate, although the degree of deficiency is less substantial than that of butyrate. Metabolic cross-feeding interactions between bacterial groups are likely important determinants of the composition of the intestinal microbial community. Acetate produced by bacterial fermentation of carbohydrates or acetogens is utilized by BPB (those using the acetyl-CoA pathway) for butyrate production, and BPB like *F. prausnitzii*, *Roseburia* spp. and *Eubacterium* spp. may grow poorly in the absence of acetate [51–53]. Thus, net acetate deficiency may contribute to deficient BPBs and butyrate and may be associated with known acetate producers such as the genera *Dorea* and *Fusicatenibacter*, which were also lower in relative abundance in ME/CFS compared to healthy controls [54, 55]. In contrast to butyrate and acetate, propionate was only reduced in patients with sr-IBS. Prior research has shown decreased concentration of fecal propionate in constipation-predominant IBS [56, 57].

Butyrate is an important health-promoting bacterial metabolite in the GI tract with diverse beneficial properties, including its role in host energy metabolism. Butyrate serves as the primary energy source for colonocytes, accounting for 70% of the energy obtained by epithelial cells in the colon [58]. In addition, butyrate influences proliferation of intestinal epithelial and stem/progenitor cells, suppresses cancer cell proliferation, and can influence epigenetic changes as a histone deacetylase inhibitor. Butyrate also plays a role in promoting epithelial barrier function by regulating HIF-1 and tight junction proteins [59].

Finally, butyrate also mediates important immunomodulatory functions in the intestine by promoting regulatory T cells, inhibiting inflammatory cytokine production, and inducing antimicrobial activity in macrophages [60].

Thus, deficiency in this intestinal homeostatic metabolite could contribute to a range of detrimental physiological disturbances including a weakened epithelial barrier and enhanced intestinal inflammation. Evidence for disruption of intestinal barrier function is supported by prior research showing elevated levels of plasma lipopolysaccharides in ME/CFS, indicative of microbial translocation [28].

### Correlation between butyrate deficiency and symptoms

The relative abundance and/or quantity of fecal BPB are inversely correlated with magnitude of symptoms, as reflected by the Multidimensional Fatigue Inventory, in ME/CFS subjects. The magnitude of general fatigue and/or physical fatigue was greater in ME/CFS subjects that had lower relative abundance or quantity of *F. prausnitzii*, *Roseburia* spp., *Ruminococcus*, and *Coprococcus*. The quantity of fecal *F. prausnitzii* based on qPCR was inversely correlated with general fatigue, physical fatigue and reduced activity. *F. prausnitzii* is an important member of the human microbiota, representing 5% of the microbiota in healthy adults. The functional importance of *F. prausnitzii* in the intestine may extend beyond its role as a BPB to include additional anti-inflammatory effects through its production of microbial anti-inflammatory molecule (MAM) as well as salicylic acid. As a potential biosensor of human health, deficiency of fecal *F. prausnitzii* has been associated with a range of other conditions including IBD, IBS, celiac disease, colorectal cancer, obesity, and more recently in patients during and even after recovery from infection with SARS-CoV-2 [61–67]. In a large cohort, the Flemish Gut Flora Project, *Faecalibacterium* along with *Coprococcus* were associated with higher quality of life scores [68]. Patients with IBD and fatigue have reduced levels of fecal *F. prausnitzii* compared with IBD patients without fatigue, further supporting the association of this bacterium with fatigue symptoms [69].

The debilitating fatigue experienced by people with ME/CFS reduces physical activity to a level substantially lower than seen in healthy controls [70–72]. In fact, individuals with ME/CFS may engage in less high-intensity physical activity than even sedentary control subjects [73]. In recent years, it has become clear from numerous studies that physical activity/exercise has a substantive effect on the composition and function of the gut microbiome. A theme is emerging in the field of exercise research based on numerous studies in laboratory animals and humans that suggests that physical activity influences the microbiome, stimulates BPB (especially *Faecalibacterium* and *Roseburia* species), and ultimately increases the levels of SCFAs [74, 75]. The mechanisms by which physical activity may influence the microbiome are not fully elucidated but such mechanisms may relate to the impact of exercise on blood flow to the intestine, gut barrier integrity, transit time in the large intestine, enterohepatic circulation of bile acids, contraction of skeletal muscles and metabolic flux, or raising of core body temperature [74, 75]. Given the debilitating nature of ME/CFS, we acknowledge that the deficiency in butyrate and BPB may arise as a result of the symptoms of ME/CFS (i.e., fatigue, post-exertional malaise, pain) and the behavioral adjustment to those symptoms (i.e., reduced physical activity). Nonetheless, even if deficient butyrate-producing capacity is a consequence rather than a direct cause of symptoms, such a deficiency could both exacerbate ME/CFS-specific symptoms or potentiate the risk for the development of additional health-related issues.

## Conclusions

Given the importance of butyrate in preventing inflammation, fortifying the intestinal barrier and promoting overall human health, the potential pathophysiological implications of butyrate deficiency in ME/CFS warrant further investigation and may provide an important target for treatment through prebiotic, probiotic or synbiotic interventions aimed at boosting butyrate production in the intestine.

## Methods

### Study Population

The initial cohort consisted of 177 ME/CFS cases and 177 healthy controls prescreened at five geographically-diverse ME/CFS clinics across the USA (Incline Village, NV; Miami, FL; New York, NY; Palo Alto, CA; Salt Lake City, UT) as part of a National Institutes of Health-sponsored R56 study. ME/CFS cases met the requirements of both the 1994 CDC [1] and the 2003 Canadian consensus criteria [9] for ME/CFS. Control participants were matched to ME/CFS cases based on geographical/clinical site, sex, age, race/ethnicity, and date of sampling (±30 days).

The CDC Criteria require that cases have fatigue persisting for greater than six months that is clinically-evaluated, persistent or relapsing, and which meets five criteria: is of new onset, is not the result of ongoing exertion, is not alleviated by rest, is made worse by exertion, and results in substantial reduction in previous levels of activity. The CDC criteria additionally require the concurrent occurrence of at least four of the following symptoms for at least six consecutive months: sore throat, tender cervical or axillary lymph nodes, muscle pain, multiple joint pain without swelling or redness, headaches of new or different type, unrefreshing sleep, post-exertional malaise, and impaired memory or concentration. The Canadian consensus criteria impose additional restrictions, requiring at least two neurologic/cognitive manifestations, and at least one clinical feature from two of the following three categories: autonomic manifestations, neuroendocrine manifestations, and immune manifestations.

Eligible cases must also have had a diminished or restricted capacity to work, reported a viral-like prodrome prior to onset of ME/CFS, and met a low-score threshold for two out of the following three domains measured by the self-reported Short Form-36 General Health Survey (SF-36): vitality <35, social functioning <62.5, role-physical <50. Additional exclusion criteria for both cases and controls included disorders or treatments resulting in immunosuppression, and antibiotic use within six weeks prior to the baseline assessment. Based on these screening criteria, we excluded five ME/CFS cases and one control participant prior to the baseline assessment.

In the current study, a nested sub-cohort was established for participants with complete survey data collection and biospecimen (stool) collection at the first and fourth (final) timepoints for the overall study. Participants were frequency-matched on key demographic elements to ensure similarity between the nested cohort and full cohort. This sub-cohort is comprised of 106 ME/CFS cases and 91 healthy controls that met these criteria; the derivation of this sub-cohort is outlined in **Supplementary Figure 6**. All subjects provided written consent in accordance with study protocols approved by the Columbia University Medical Center Institutional Review Board (IRB).

### Clinical Assessments

All subjects completed standardized screening instruments to assess medical history, family medical history, current medication use, symptom scores, and demographic/lifestyle information, as well as a baseline SF-36. All subjects underwent a screening blood draw to determine that they had normal values in the following three laboratory tests from Quest Diagnostics: complete blood count with differential, comprehensive metabolic panel, thyroid stimulating hormone (TSH).

Subjects eligible for participation after the baseline visit returned to the same clinic four times over the course of one year for sample collection and completion of survey instruments. At each of the four visits subjects completed a range of rating scales, including the Multidimensional Fatigue Inventory (MFI). Prior to all four study visits, subjects were provided with at-home collection kits for stool. Stool samples were collected within 48 hours prior to each study visit and refrigerated until the day of the visit. All samples were shipped to the Columbia University laboratory site in insulated Styrofoam boxes with frozen and refrigerated gel packs. The stool samples were aliquoted, weighed and moved to -80 °C freezers for storage. Only samples and clinical rating scale data obtained at the first study visit were analyzed as part of the present study.

On the medical history questionnaire, participants were asked to self-report if they received an IBS diagnosis (sr-IBS) by a physician and the date of diagnosis. In the analytic sub-cohort, 35 out of the 106 (33.0%) ME/CFS cases reported sr-IBS, while only 3 out of the 91 control subjects (3.3%) reported sr-IBS. The use of probiotic and prebiotic supplements was specifically included in the “current medication use” data collection instrument, which determined the frequency and recent use of these products. Any subjects that indicated consumption of these supplements daily or a few times a week and reported using them within the last week prior to their study visit would be endorsed for that type of supplement. Participants also reported any antibiotic use that occurred outside of the six-week window for study eligibility.

The MFI consists of a 20-item self-reported questionnaire that evaluates five dimensions of fatigue: general, physical and mental fatigue, reduced activity, and reduced motivation [76]. The scoring for this instrument was transformed into a 0–100 scale to allow for comparisons between dimensions: a score of 100 was equivalent to maximum disability or severity and a score of zero was equivalent to no disability or disturbance.

### Fecal DNA Extraction

A modified protocol of the QIAmp DNA Stool Mini Kit (Qiagen Inc; Valencia CA, USA) was used for the extraction of DNA from stool samples. Prior to the initiation of the extraction process, all disposable elements including tubes, columns, 0.1mm and 0.5 mm glass beads (MoBio Laboratories) were UV irradiated twice at a distance of 1 inch from UV bulbs and at 3000 x 100 μJ/cm^2^ in a SpectroLinker XL-1500 UV crosslinker (Spectronics Corporation). All liquid extraction reagents in the kit were aliquoted into 2 mL tubes in a UV hood at ≤1 ml and UV irradiated as mentioned above. To confirm that bacterial 16S rDNA contaminants were not introduced from the extraction reagents, two empty 2 mL tubes were used for each set of DNA extraction to serve as negative controls throughout the entire process. Each fecal sample was weighed and < 220 mg of each fecal sample was re-suspended in AL Buffer (Qiagen). Glass beads (0.1mm and 0.5mm, MoBio Laboratories) were added to the mixture. To ensure Gram-positive bacteria lysis, samples were disrupted by bead beating in a TissueLyser (Qiagen) for 5 minutes at 30Hz and incubated for 5 minutes at 95°C. The remaining steps for DNA extraction were performed in a UV hood by following the manufacturer’s protocol. To determine DNA concentration and purity, a NanoDrop ND-100 spectrophotometer (NanoDrop Technnologies, Wilmington, DE) was used and the extracted DNA samples were stored at -80°C.

### Shotgun metagenomic sequencing and bioinformatic analyses

Shot-gun metagenomics sequencing was carried out on DNA extracts obtained from 197 fecal samples (106 ME/CFS cases and 91 healthy controls). For Illumina library preparation, genomic DNA was sheared to a 200-bp average fragment length using a Covaris E210 focused ultrasonicator. Sheared DNA was purified and used for Illumina library construction using the KAPA Hyper Prep kit (KK8504, Kapa Biosystems). Sequencing libraries were quantified using an Agilent Bioanalyzer 2100. Sequencing was carried out on the Illumina HiSeq 4000 platform (Illumina, San Diego, CA, USA). Sequencing libraries from samples of cases and controls were grouped into eight different sequencing pools. Each sample yielded 27.8 ± 10.3 (mean ± sd) million reads. The demultiplexed raw FastQ files were adapter trimmed using Cutadapt [77]. Adaptor trimming was followed by the generation of quality reports using FastQC and filtering with PrinSEQ [78]. Host background levels were determined by mapping the filtered reads against the human genome using Bowtie2 mapper [79]. After the step of host subtraction, 25.1 ± 9.0 (mean ± sd) million reads per sample remained on average. Non-host reads were subjected to Kraken2 for taxonomy classification [80]. Kraken2 matches each K-mer within a query sequence to the lowest common ancestor (LCA) of all genomes in the database containing the given K-mer. Our Kraken2 local database included all 16,799 fully sequenced and representative bacteria species genomes in the RefSeq database (December 2018). The species-level taxonomy abundances were estimated using Bracken, which is recommended to perform a Bayesian estimation of taxonomy abundance after the use of Kraken2 [80, 81]. Structural zeros in the abundance table were further identified using the program Analysis of Microbiome Data in the Presence of Excess Zeros version II (ANCOM-II) [82]. The three groups we have compared in our study include ME/CFS with sr-IBS, ME/CFS without sr-IBS, and healthy controls. The taxa presenting as structural zeros in all three categories were eliminated from the dataset. The data was rarefied prior to diversity analyses using a depth of 500,000 reads. Diversity metrics (alpha diversity: Shannon index, pielou’s evenness, and observed otus; beta-diversity: Bray-Curtis dissimilarity) were calculated and plotted using the core-diversity plugin and the emperor plugin within QIIME2 [83]. The beta diversity significance among groups was examined by QIIME2 diversity plugin with PERMANOVA tests. To evaluate fecal metabolic functional profiles from SMS data, KEGG gene family abundances and the functional pathway and module abundances were calculated using FMAP with the default database [84]. The abundance of genes involved in butyrate synthesis was calculated by aligning host subtracted sequence reads to a curated database specifically consisting of butyrate synthesis pathway related genes [41], using customized scripts from the FMAP pipeline.

### qPCR

Plasmid DNA standards were constructed using published qPCR primers targeting the bacterial 16S rRNA gene of *Faecalibacterium prausnitzii* and *Eubacterium*/*Roseburia* genera [85], the bacterial *but* gene (BcoA) [42], and broad range bacterial 16S rRNA genes to quantitate total bacteria [86] for absolute quantitation. Bacterial gene targets were first amplified by conventional PCR from human stool samples. PCR products were run on 1% agarose gels and purified using the QIAgen Gel Extraction kit. Purified products were ligated into pGEM T-Easy vector (Promega), transformed into DH5α competent cells, and cultured on Luria Bertani plates with ampicillin. Colonies were inoculated into Luria broth with ampicillin (5 ml), and plasmids were extracted with Pure Link Plasmid Extraction Kit (Invitrogen). After verifying that plasmid sequences had 100% nucleotide similarity to the target bacterial taxa or the bacterial *but* gene, plasmids were linearized with the restriction enzyme SphI, and 10-fold serial dilutions were generated ranging from 5 x 10^6^ to 5 copies (note for *Eubacterium*/*Roseburia* standards, two plasmids were combined consisting of one clone matching *Eubacterium hallii* and one clone matching *Roseburia hominis*). Salmon sperm DNA at 2.5 ng/μl was spiked into the standard dilutions as background DNA. Real time PCR was performed with the ABI StepOne Plus Real Time PCR system (Applied Biosystems). Amplification plots for all plasmid standards were sensitive down to five copies and standard curves generated yielded correlation coefficients ranging from 0.998 to 1. For probe-based assays (total bacteria), each 25 μL real time PCR reaction consisted of 1x TaqMan Universal PCR Master Mix (Applied Biosystems), 300 nM primers, 200 nM probe and 5 ng of fecal DNA. The total bacteria probe was labeled with a 5’-end fluorescent reporter (6-carboxyfluorescine, FAM) and had a 3’-end black hole quencher (BHQ1a, Operon). For SYBR green-based assays, each 25 μl reaction consisted of 1X SYBR Green PCR master mix (Applied biosystems), 300 nM primers and 5ng of fecal DNA. Cycling conditions consisted of 50**°**C for 2 min, 95**°** C for 10 min, and 45 cycles at 95**°**C for 15 s and 60**°**C for 1 min 30 s (for total bacteria, *Eubacterium*/*Roseburia* 16s *and Faecalibacterium* 16s assays). For *but* gene the cycle conditions were 50**°**C for 2 min, 95**°** C for 10 min, and 45 cycles at 95**°**C for 15 s, 53**°**C for 1 min 45 s and 77**°**C for 30 s (data collection). All samples were run in duplicate and averaged for each assay. Absolute quantity was determined based on the weight of fecal samples and expressed as copy numbers per gram of feces. The calculated relative abundances of bacterial taxa and the *but* gene were also assessed by dividing the absolute copy number for the targeted bacteria (*F. prausnitzii* or *Eubacterium*/*Roseburia*) or *but* gene by the total bacterial 16S copy number.

### Fecal SCFA metabolomics

Fecal SCFAs were measured by gas chromatography-mass spectrometry (GC-MS) as previously described [87] with minor modifications. Fecal samples were homogenized and 10 mg resuspended in 0.5 ml 2-ethylbutyric acid (100 μg/ml; as internal standard 1) in LC/MS grade water. The sample was acidified with 0.1 ml concentrated hydrochloric acid, vortexed and placed on ice. 1 ml of 4-methylvaleric acid (100 μg/ml; as internal standard 2) in tert-butyl methyl ether (MTBE) was added, the sample was vortexed and shaken for 30 min at room temperature, and centrifuged for 2 min at 14,000 rcf. Supernatant was collected after centrifugation and 0.1 g of anhydrous sodium sulfate was added to it, vortexed and incubated at room temperature for 10 min. Finally, 100 μl of the supernatant was combined with 25 μl N-tert-Butyldimethylsilyl-N-methyltrifluoroacetamide (MTBSTFA). Calibration standards containing acetic acid, n-butyric acid, and propionic acid as well as the internal standards, 2-ethylbutyric acid and 4-methylvaleric acid, were prepared and derivatized with MTBSTFA. After adding MTBSTFA as a derivatization reagent and incubating for 30 min at 80°C followed by 1 hour at room temperature, samples were injected with a 100:1 split into an Agilent GC7890B gas chromatograph coupled with an Agilent 5977MS mass spectrometer detector. Helium was used as a carrier gas. Analyses were performed using a DB5 duraguard capillary column (30 m x 0.25 mm x 0.25 μm film thickness). The column head pressure was 9.83 psi. Injector, source and quadrupole temperatures were 250°C, 290°C, 150°C, respectively. The GC oven program was 50°C for 0.5 min, increased to 70°C for 3.5 min at 5°C/min, increased to 120°C at 10°C/min, and increased to 290°C for 3 min at 35°C/min. The total run time was 20.857 min.

### Statistical analyses

#### Cohort characteristics

Differences in cohort characteristics between ME/CFS and healthy control subjects were assessed using the Mann-Whitney U test for continuous variables and either the Fisher’s Exact test or Chi-squared test for categorical variables and p < 0.05 was considered statistically significant.

#### Box-and-whiskers plots, Mann–Whitney U-test and Kruskal-Wallis test

All box-and-whiskers plots shown in the main and supplementary figures represent the interquartile ranges (25th through 75th percentiles, boxes), medians (50th percentiles, bars within the boxes), the 5th and 95th percentiles (whiskers above and below the boxes), and outliers beyond the whiskers (closed circles). Plots were created and statistical analyses performed using Prism 7 (GraphPad Software, CA). All statistics based on data presented in box-and-whiskers plots comparing all ME/CFS subjects and healthy controls are two-tailed *P-*values derived from Mann–Whitney *U*-tests. For stratified analyses of more than two groups, significance was first determined based on the Kruskal-Wallis test. If the Kruskal-Wallis test was significant at p < 0.05, then between group significance was determined based on the Mann-Whitney U test with multiple testing (Bonferroni) correction applied, and an adjusted p-value (p^adj^) < 0.05 was considered significant.

#### Beta diversity

Beta diversity was assessed based on the Bray-Curtis dissimilarity metric. Differences in beta diversity using Principal coordinate analyses (PCoA) were visualized with 3-D plots using QIIME2. Permutational multivariate analysis of variance (PERMANOVA) with 999 Monte Carlo permutations was used to evaluate the statistical significance of the dissimilarity between groups. The Freedman-Lane PERMANOVA test [88] was used to assess differences in microbiota beta diversity between ME/CFS and healthy controls, and was adjusted for covariates (sr-IBS status, site of sampling, sex, BMI, race/ethnicity, age, antibiotic usage 6-12 weeks prior to sample collection, probiotic supplement use, and prebiotic supplement use). Two-group comparisons were considered significant at p < 0.05. For stratified analyses (more than two groups), the FDR was controlled by Bonferroni correction and an adjusted p-value (p^adj^) < 0.05 was considered significant.

#### Differential Taxa Abundance, GAMLSS-BEZI models

Prior to differential abundance analysis, PERFect:Permutation was used to remove taxa that are present due to contamination or otherwise are unrelated to ME/CFS [89]. The regression then used to analyze the binary outcome ME/CFS was a Generalized Additive Model for Location, Scale and Shape with a zero-inflated beta distribution (GAMLSS-BEZI) performed using the metamicrobiomeR package [39]. This package was built in order to deal with zero inflated, compositional data such as microbiome relative abundance. Features in the abundance table were pre-filtered, using the built-in pre-filtering step in metamicrobiomeR, to remove features with mean relative abundance ≤ 0.1% and with prevalence ≤ 5% across samples. With GAMLSS-BEZI a multivariate regression was performed with ME/CFS status as the variable for comparison and adjusted for covariates (sr-IBS diagnosis, site of recruitment, sex, BMI, race and ethnicity, age, antibiotic use within 6-12 weeks of testing, probiotic supplement use and prebiotic supplement use) for each taxon (species and genera were assessed separately) present in the data. Use of prescription narcotics and antidepressants were tested in each model but did not affect the results and were therefore not included in the final models. Multi-level categorical variables were assigned as dummy variables and continuous variables were scaled using z-scores. The original dataset includes 197 observations, 106 cases and 91 controls. Four models were assessed, Model 1: comparing ME/CFS vs. healthy controls, Model 2: ME/CFS without sr-IBS vs. healthy controls without sr-IBS, Model 3: ME/CFS with sr-IBS vs. healthy controls without sr-IBS, and Model 4: ME/CFS with sr-IBS vs. ME/CFS without sr-IBS. The *p values* were adjusted for multiple comparisons using Benjamini-Hochberg false discovery rate (FDR) procedure, and adjusted *p value* < 0.05 was considered statistically significant. The odds ratio and 95% confidence intervals are used to interpret the model.

#### Differential gut bacterial metabolic pathway modules

KEGG ontology profiles, determined using FMAP, were assigned to gut metabolic module profiles using GOmixer [40]. Analyses focused on the top 2000 most abundant bacterial KEGG orthologs. Statistically over/under-represented gut metabolic modules between ME/CFS and controls were determined using GOmixer, which applies a Wilcoxon rank-sum test and the Benjamini-Hochberg false discovery rate (FDR) to correct for multiple testing. Data were scaled and the mean differences in pathways and modules were considered significant at an FDR adjusted p-value < 0.1.

#### Regression analyses

Fecal bacterial alpha diversity, qPCR quantities of fecal bacteria and the *but* gene, fecal SCFAs concentrations and metagenomic content of genes involved in butyrate synthesis pathways were all assessed with generalized linear regressions. All models were adjusted for covariates (sr-IBS diagnosis, testing site, sex, BMI, race and ethnicity, age, antibiotic use within 6-12 weeks of testing, probiotic supplement use and prebiotic supplement use). Use of prescription narcotics and antidepressants were tested in each model but did not affect the results and were therefore not included in the final models. Data distribution was evaluated using histograms for each outcome, and a variety of linear models were chosen after model fit was tested utilizing the BIC measure. For count data a negative binomial and Poisson regression were evaluated, for continuous data with a correct assumption of a normal distribution a linear regression was used, and for continuous data with a non-normal distribution a generalized linear regression with Gamma distribution with log link model was compared with a lognormal generalized linear regression model. The SCFA concentrations were assessed with a generalized linear regression with Gamma distribution with log link. The alpha diversity measures of Shannon index and Evenness were log transformed and were evaluated with a linear regression assuming normal distribution. The observed species measure was assessed using a negative binomial generalized linear model. The metagenomic content for each butyrate-producing gene in counts per million (CPM) was estimated with a generalized linear regression with Gamma distribution with log link. Finally, the RT-qPCR data in copies/gram feces was evaluated with a generalized linear regression with Gamma distribution with log link. For the generalized linear models with Gamma distribution with log link and the negative binomial generalized linear model, the exponent of the estimate is the fold change, and the coefficients of linear regressions assuming normal distribution indicates the linearity of the association between the predictors and outcome. For each regression an interaction term between ME/CFS and sr-IBS was tested to determine if the relationship between ME/CFS and the outcome was altered significantly by sr-IBS status. When the interaction term was not significant, it was removed from the final models. The interaction term was only significant for SCFAs models, and is shown in the result. Stratified analyses were conducted to evaluate the nature of the interaction in these models. Results were considered significant for predictor variables for all regression models when p-value < 0.05.

#### Correlations

Correlations among measured variables (alpha diversity, relative abundance of bacterial taxa, qPCR quantity of bacterial taxa and the bacterial *but* gene, metagenomic content for genes along the acetyl-CoA pathway of butyrate production, and the concentration of fecal SCFAs) and between measured variables and the scores for the five dimensions (General Fatigue, Physical Fatigue, Mental Fatigue, Reduced Activity, and Reduced Motivation) of the Multidimensional Fatigue Inventory (MFI) were examined using nonparametric Spearman’s correlation. FDR was controlled with the Benjamini-Hochberg procedure. Correlations with adjusted p-values < 0.05 were considered significant. Correlograms were created using the corrplot package in R.

## Data Availability

Host subtracted, shotgun metagenomic sequences from fecal samples of all ME/CFS and healthy control samples are available from the Sequence Read Archive (SRA) under PRJNA751448, which will be made publicly available upon peer reviewed publication of this manuscript. All additional relevant data are available in this article and its Supplementary Information files, or from the corresponding author upon request.

## Declarations

### Ethics approval and consent to participate

All subjects provided written consent in accordance with study protocols approved by the Columbia University Medical Center Institutional Review Board (IRB).

### Consent for publication

Not applicable.

### Availability of data and material

Host subtracted, shotgun metagenomic sequences from fecal samples of all ME/CFS and healthy control samples are available from the Sequence Read Archive (SRA) under PRJNA751448. All additional relevant data are available in this article and its Supplementary Information files, or from the corresponding author upon request.

### Author Contributions

WIL, TB, AC, DM, MH, ALK, SL, LB, SDV, NGK, JGM, DLP, and BLW contributed to study design. WIL, TB, OA, AR, and BLW designed the experiments. CG, XC, OA, RAY, AC, AR, and BLW designed analyses. CG, XC, OA, RAY, AR, and BLW analyzed the data. CG, XC, RAY, AC and BLW wrote the paper. All authors reviewed and edited the manuscript.

## Acknowledgements

Not applicable

## Supplementary Figures and Tables

**Supplementary Figure 1:**
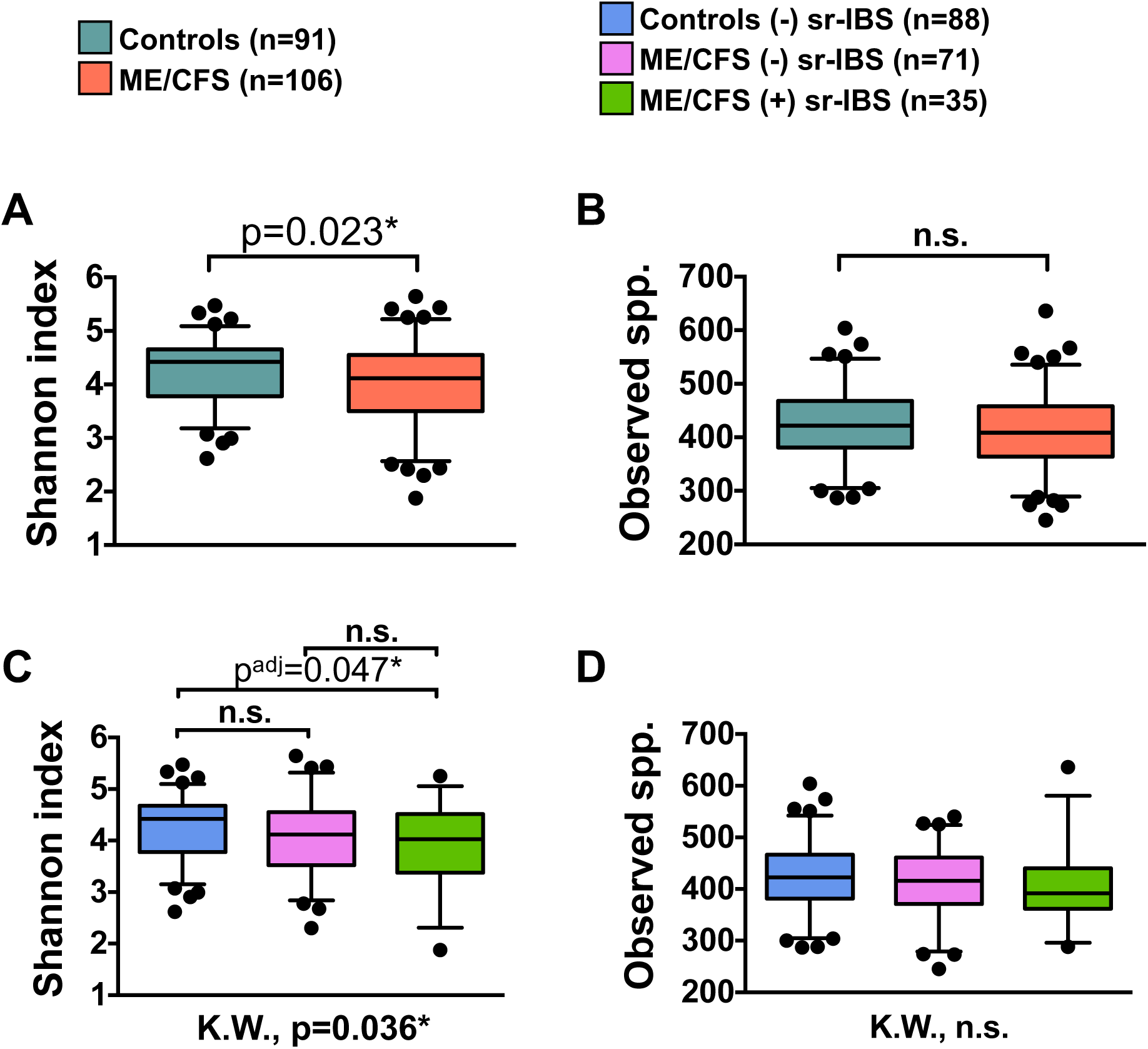
Gut microbiome alpha diversity in ME/CFS. Box-and-whiskers plots showing the distribution of microbiome alpha diversity (Shannon diversity index and Observed species) between ME/CFS and healthy controls (**A, B**) and among stratified groups for healthy controls without (-) sr-IBS, ME/CFS subjects without (-) sr-IBS, and ME/CFS subjects with (+) sr-IBS (**C, D**). Statistical significance was determined based on two-tailed p-values from the Mann-Whitney U test (**A, B**). For stratified analyses significance was first determined based on the Kruskal-Wallis test (K.W., results shown below each figure in **C, D**). If significant (p < 0.05) based on K.W., then between group significance was determined based on the Mann-Whitney U test with multiple testing (Bonferroni) correction (p^adj^-value). n.s. = not significant; * = p or p^adj^ < 0.05.

**Supplementary Figure 2:**
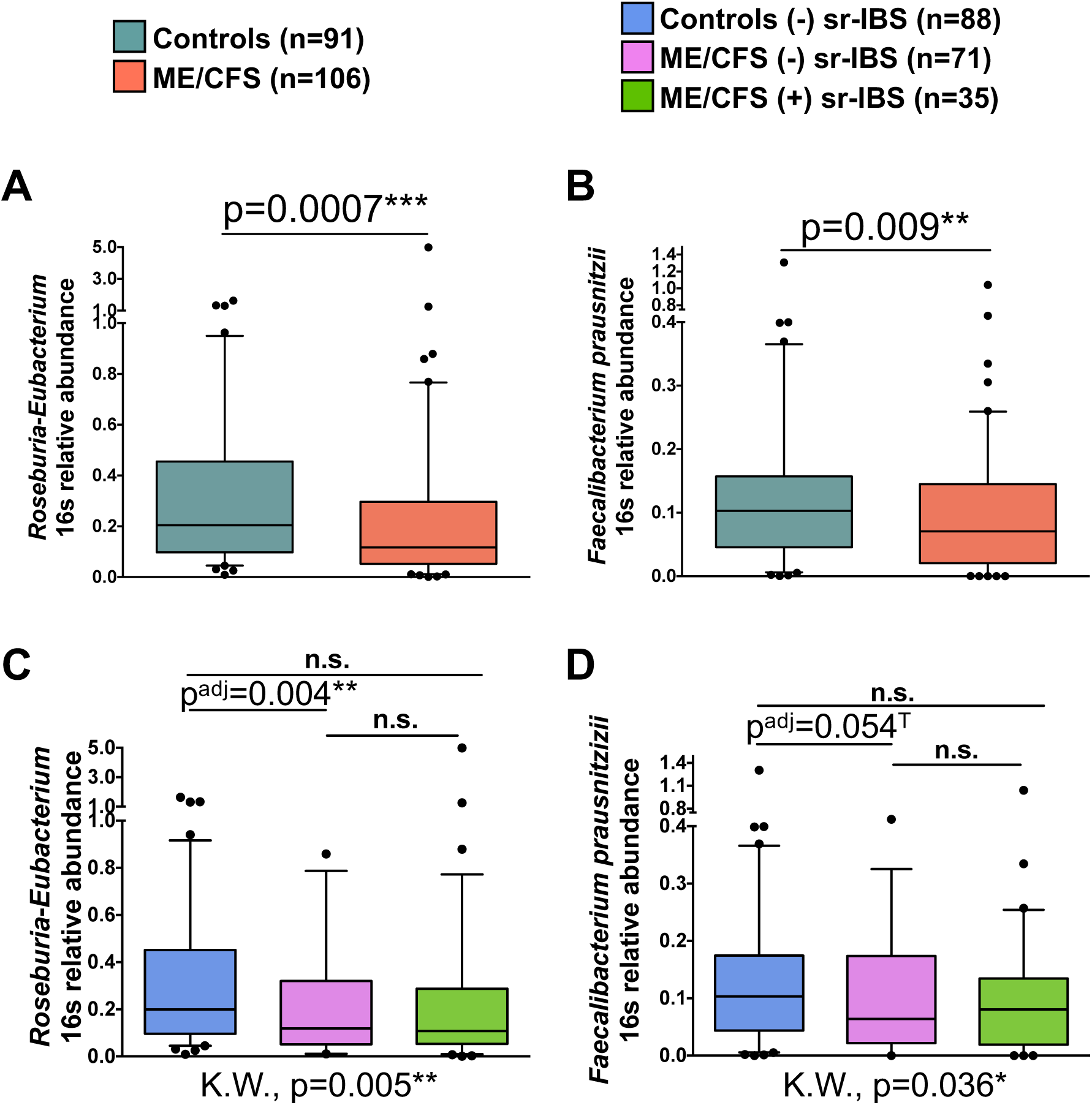
Calculated relative abundance of butyrate producing taxa in feces based on qPCR. Box-and-whiskers plots showing the distribution of calculated relative abundance for fecal *Roseburia-Eubacterium* 16S rRNA and *F. prausnitzii* 16S rRNA between ME/CFS and healthy controls (**A, B**) and among stratified groups for healthy controls without (-) sr-IBS, ME/CFS subjects without (-) sr-IBS, and ME/CFS subjects with (+) sr-IBS (**C, D**). Statistical significance was determined based on two-tailed p-values from the Mann-Whitney U test (**A, B**). For stratified analyses significance was first determined based on the Kruskal-Wallis test (K.W., results shown below each figure in **D-F**). If significant (p < 0.05) based on K.W., then between group significance was determined based on the Mann-Whitney U test with multiple testing (Bonferroni) correction (p^adj^-value). n.s. = not significant; * = p or p^adj^ < 0.05; ** = p or p^adj^ < 0.01; *** = p or p^adj^ < 0.001; T = trend (p or p^adj^ < 0.1).

**Supplementary Figure 3:**
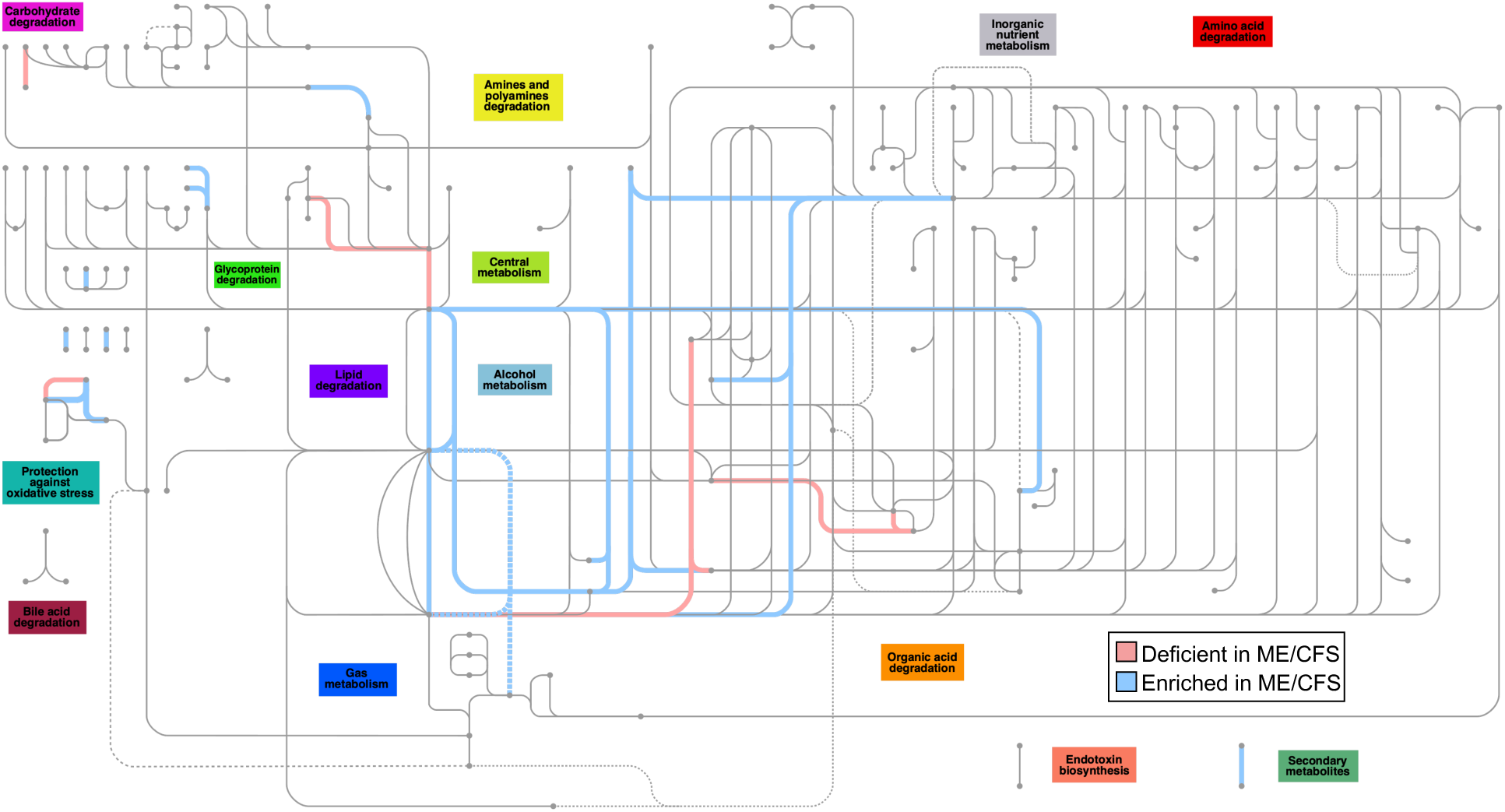
Gut microbiome module map showing deficient and enriched modules in ME/CFS. In the map, microbiome modules that are deficient in ME/CFS relative to healthy controls are shown as red highlighted connections. Modules that are enriched in ME/CFS relative to healthy controls are shown as blue highlighted connections.

**Supplementary Figure 4:**
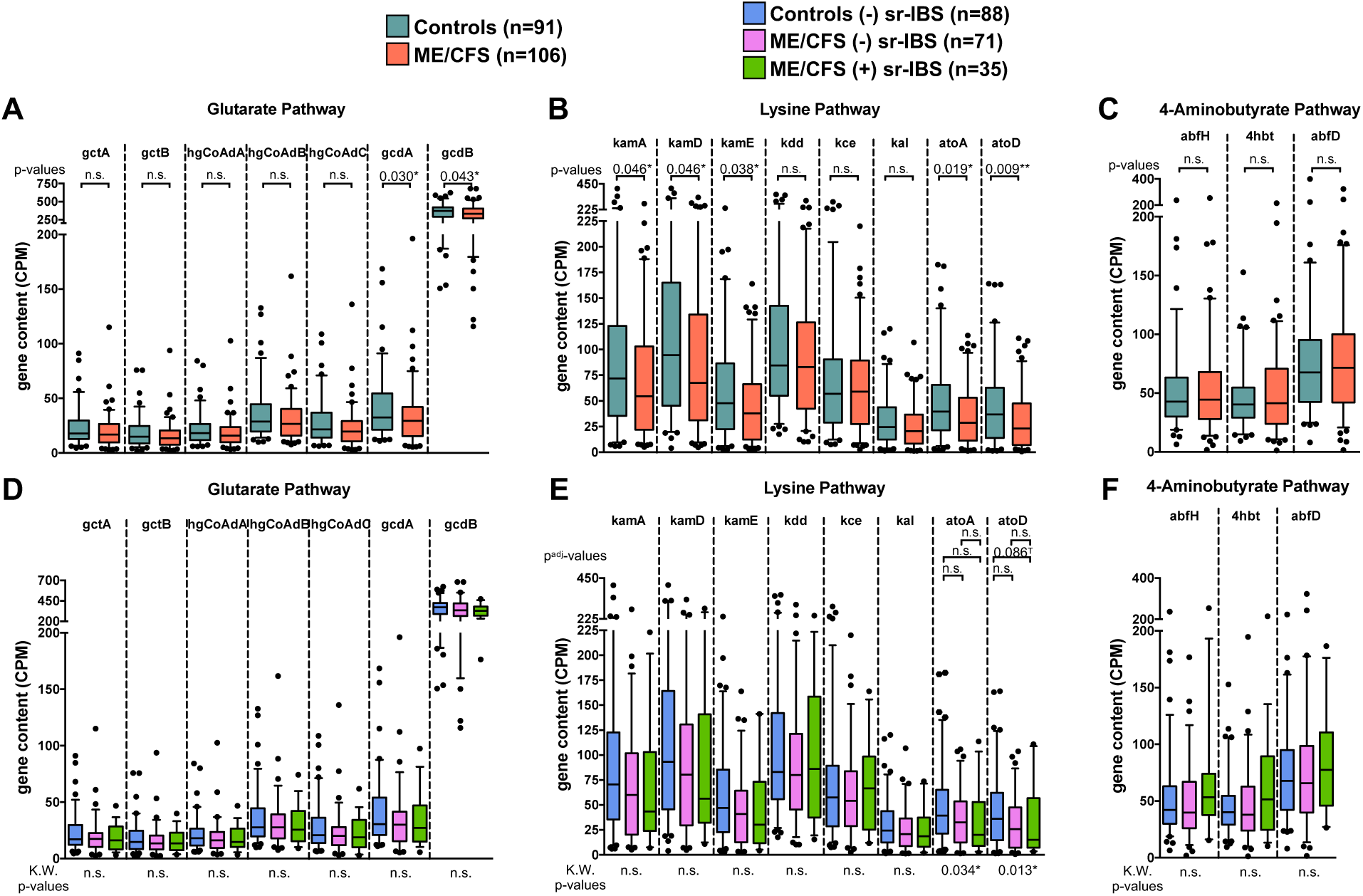
Metagenomic gene content of the fecal microbiota for genes in bacterial pathways of butyrate production. Box-and-whiskers plots showing the distribution of gene counts (CPM) for genes in the Glutarate (**A, D**), Lysine (**B, E**), and 4- Aminobutyrate (**C, F**) pathways of butyrate production between ME/CFS and healthy controls (**A-C**) and among stratified groups for healthy controls without (-) sr-IBS, ME/CFS subjects without (-) sr-IBS, and ME/CFS subjects with (+) sr-IBS (**D-F**). Statistical significance was determined based on two-tailed p-values from the Mann-Whitney U test for each gene (**A-C**). For stratified analyses significance was first determined based on the Kruskal-Wallis test for each gene (K.W., results shown below each figure in **D-F**). If significant (p < 0.05) based on K.W., then between group significance was determined based on the Mann-Whitney U test with multiple testing (Bonferroni) correction (p^adj^-value). n.s. = not significant; * = p or p^adj^ < 0.05; ** = p or p^adj^ < 0.01; T = trend (p or p^adj^ < 0.1).

**Supplementary Figure 5:**
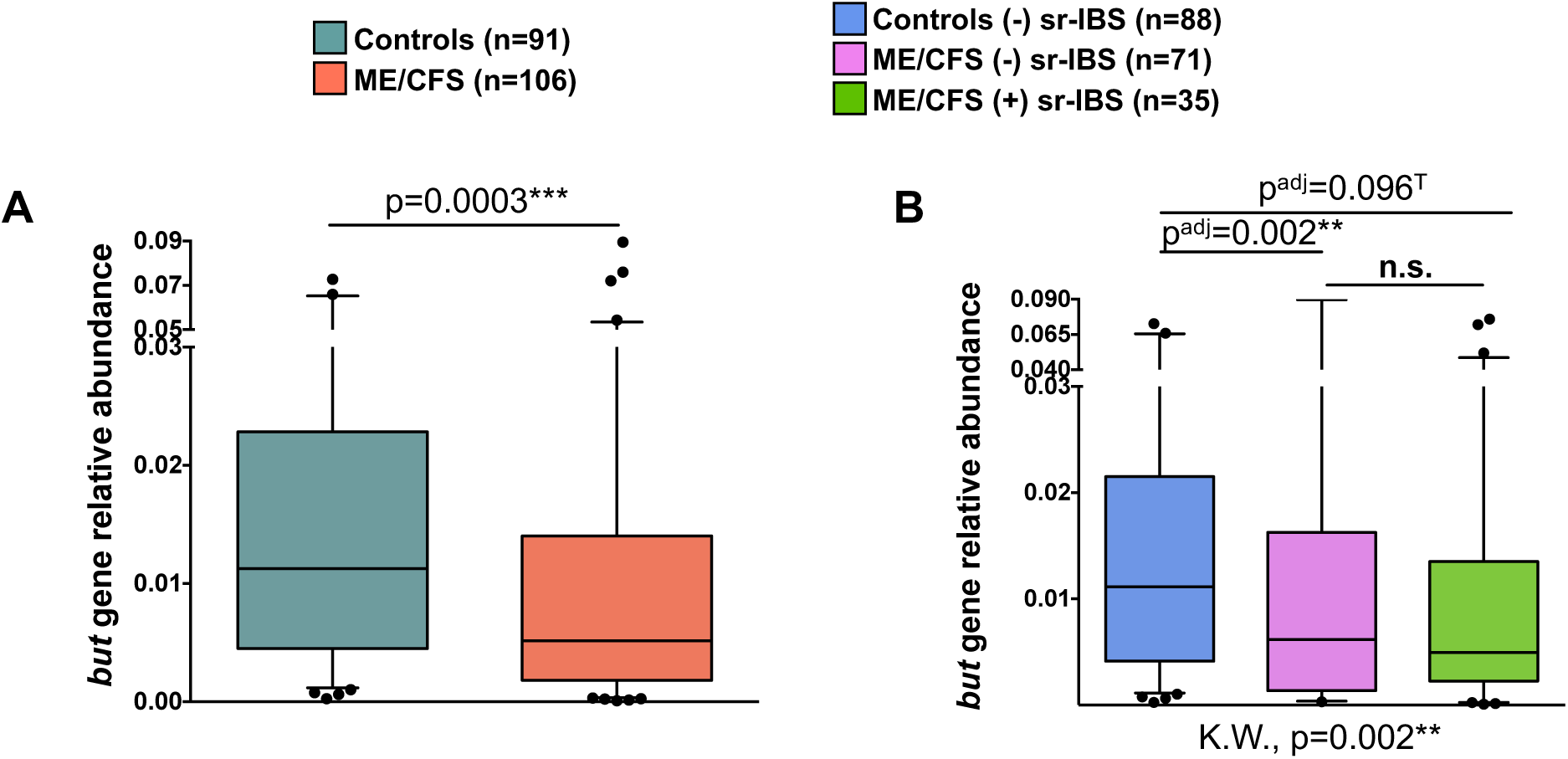
Calculated relative abundance of bacterial *but* gene in feces based on qPCR. Box-and-whiskers plots showing the distribution of calculated relative abundance for fecal *but* gene between ME/CFS and healthy controls (**A**) and among stratified groups for healthy controls without (-) sr-IBS, ME/CFS subjects without (-) sr-IBS, and ME/CFS subjects with (+) sr-IBS (**B**). Statistical significance was determined based on two-tailed p-values from the Mann-Whitney U test (**A**). For stratified analyses significance was first determined based on the Kruskal-Wallis test (K.W., results shown below **B**). If significant (p < 0.05) based on K.W., then between group significance was determined based on the Mann-Whitney U test with multiple testing (Bonferroni) correction (p^adj^-value). n.s. = not significant; ** = p or p^adj^ < 0.01; *** = p or p^adj^ < 0.001; T = trend (p or p^adj^ < 0.1).

**Supplementary Figure 6:**
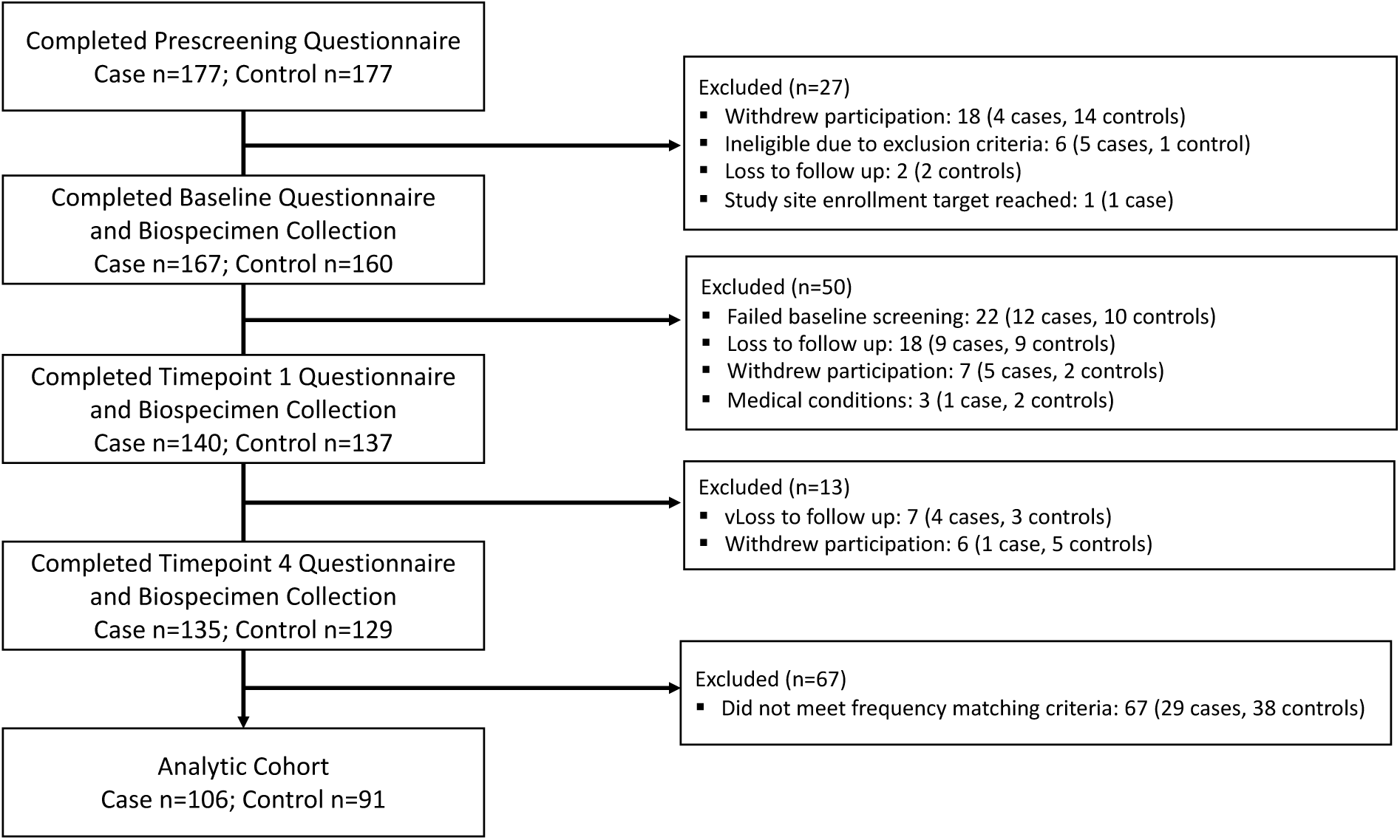
Flowchart for cohort sample selection.

**Supplementary Table 1A:**
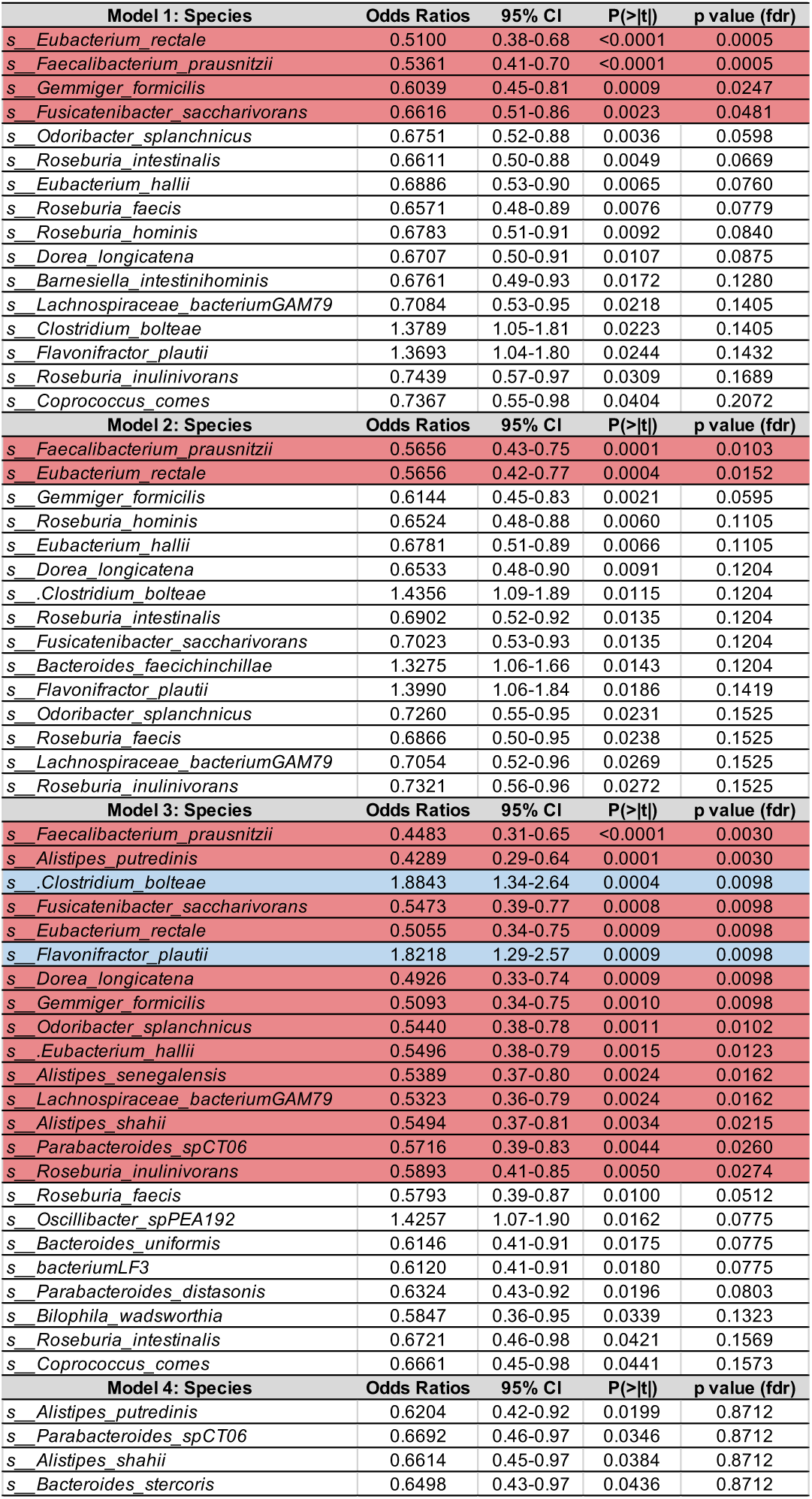
Species-level results from GAMLSS-BEZI models for differential abundance of species between: Model 1: All ME/CFS vs healthy controls, Model 2: ME/CFS without sr-IBS vs healthy controls without sr-IBS, Model 3: ME/CFS with sr-IBS vs healthy controls without sr-IBS, and Model 4: ME/CFS without sr-IBS vs ME/CFS with sr-IBS. Models are adjusted for sr-IBS diagnosis (only model 1), site of recruitment, sex, BMI, race and ethnicity, age, antibiotic use within 6-12 weeks of sample collection, probiotic supplement use, and prebiotic supplement use. All species with significant unadjusted p-values are shown for each model. Species highlighted in red are significantly decreased in the ME/CFS groups after FDR adjustment. Species highlighted in blue are significantly increased in the ME/CFS group after FDR adjustment. Odds ratios, 95% CI, unadjusted p-values (P(>|t|), and FDR adjusted p-values are shown.

**Supplementary Table 1B.**
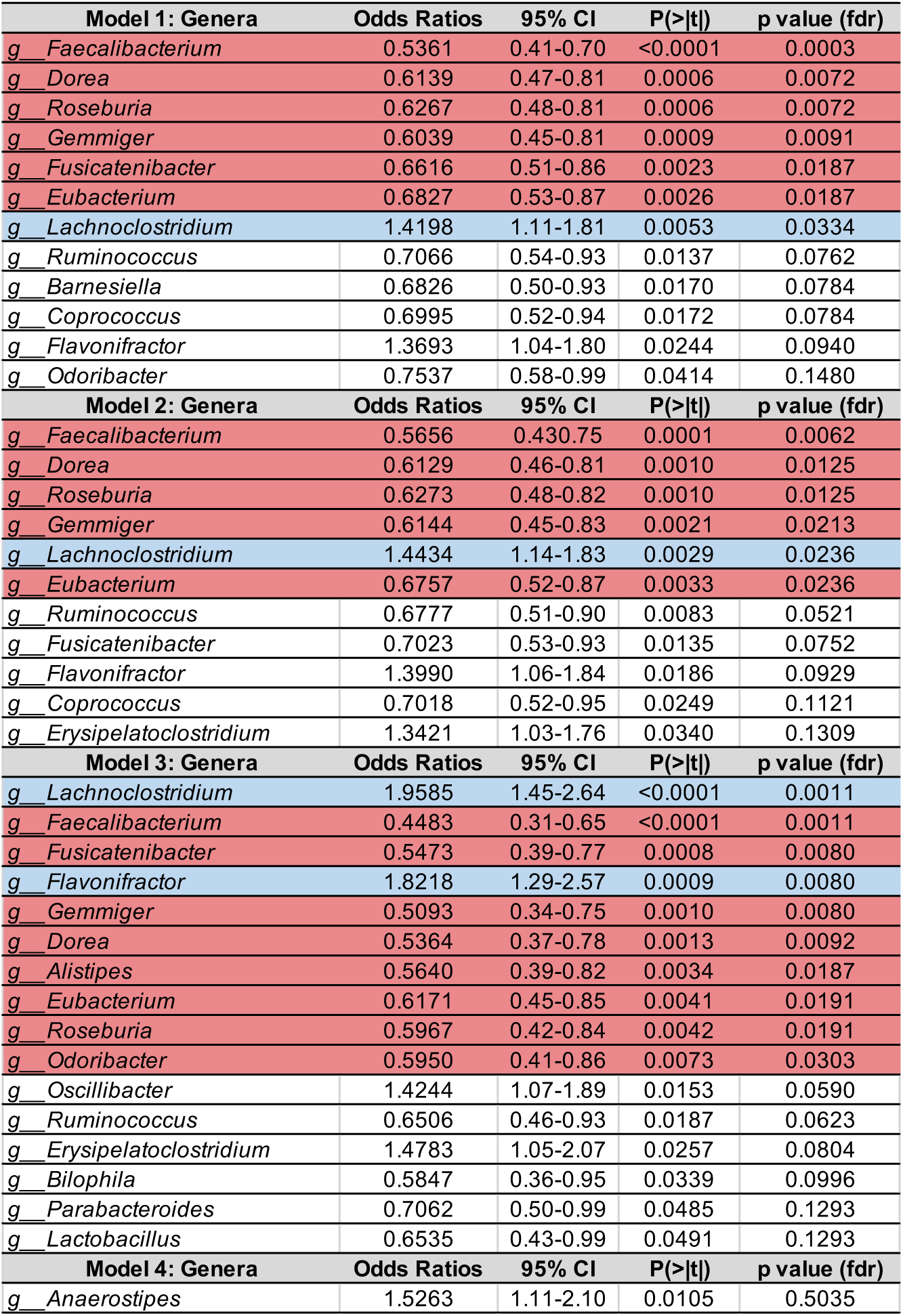
Genus-level results from GAMLSS-BEZI models for differential abundance of genera between: Model 1: All ME/CFS vs healthy controls, Model 2: ME/CFS without sr-IBS vs healthy controls without sr-IBS, Model 3: ME/CFS with sr-IBS vs healthy controls without sr-IBS, and Model 4: ME/CFS without sr-IBS vs ME/CFS with sr-IBS. Models are adjusted for sr-IBS diagnosis (only model 1), site of recruitment, sex, BMI, race and ethnicity, age, antibiotic use within 6-12 weeks of sample collection, probiotic supplement use, and prebiotic supplement use. All species with significant unadjusted p-values are shown for each model. Species highlighted in red are significantly decreased in the ME/CFS groups after FDR adjustment. Species highlighted in blue are significantly increased in the ME/CFS group after FDR adjustment. Odds ratios, 95% CI, unadjusted p-values (P(>|t|), and FDR adjusted p-values are shown.

**Supplementary Table 2:** Generalized linear regression showing the relationship between ME/CFS status and the calculated relative abundance based on qPCR of *Roseburia/Eubacterium* taxa, *F. prausnitzii* and the bacterial *but* gene.

| *Predictors* | *Roseburia/Eubacterium (calculated relative abundance)* |  |  | *F. prausnitzii (calculated relative abundance)* |  |  | *but gene (calculated relative abundance)* |  |  |
| --- | --- | --- | --- | --- | --- | --- | --- | --- | --- |
|  | *Fold Change* | *95% CI* | *p* | *Fold Change* | *95% CI* | *p* | *Fold Change* | *95% CI* | *p* |
| (Intercept) | 0.39 | 0.22 – 0.68 | **0.001** | 0.24 | 0.14 - 0.40 | **<0.001** | 0.02 | 0.01 - 0.03 | **<0.001** |
| ME/CFS | 0.62 | 0.42 – 0.92 | **0.019** | 0.60 | 0.42 - 0.85 | **0.004** | 0.60 | 0.39 - 0.91 | **0.017** |
| sr-IBS | 1.13 | 0.71 – 1.80 | 0.607 | 1.03 | 0.68 - 1.56 | 0.877 | 1.22 | 0.74 - 2.02 | 0.435 |
| Site 2(Florida) | 1.24 | 0.62 – 2.47 | 0.547 | 0.80 | 0.43 - 1.48 | 0.483 | 1.64 | 0.78 - 3.48 | 0.196 |
| Site 3(Nevada) | 0.86 | 0.48 – 1.55 | 0.618 | 0.70 | 0.42 - 1.18 | 0.182 | 1.21 | 0.64 - 2.29 | 0.551 |
| Site 4(New York) | 1.29 | 0.75 – 2.24 | 0.357 | 0.87 | 0.53 - 1.42 | 0.574 | 1.04 | 0.57 - 1.88 | 0.900 |
| Site 5(Utah) | 0.90 | 0.54 – 1.50 | 0.685 | 0.79 | 0.5 - 1.24 | 0.302 | 1.15 | 0.66 - 1.99 | 0.633 |
| Sex | 0.89 | 0.58 – 1.37 | 0.595 | 0.59 | 0.4 - 0.86 | **0.007** | 0.94 | 0.59 - 1.51 | 0.807 |
| BMI | 0.99 | 0.83 – 1.18 | 0.890 | 0.85 | 0.72 - 0.99 | **0.035** | 0.91 | 0.75 - 1.1 | 0.319 |
| Hispanic | 0.39 | 0.17 – 0.90 | **0.030** | 0.62 | 0.29 - 1.32 | 0.215 | 0.30 | 0.12 - 0.74 | **0.010** |
| Non-White & Non-Hispanic | 1.09 | 0.51 – 2.33 | 0.831 | 0.97 | 0.49 - 1.92 | 0.940 | 1.67 | 0.73 - 3.82 | 0.225 |
| Age | 0.89 | 0.74 – 1.07 | 0.234 | 0.98 | 0.83 - 1.16 | 0.822 | 1.00 | 0.82 - 1.22 | 0.998 |
| Antibiotics | 1.99 | 1.09 – 3.63 | **0.026** | 1.25 | 0.73 - 2.13 | 0.424 | 1.29 | 0.67 - 2.48 | 0.448 |
| Probiotics | 0.82 | 0.54 – 1.25 | 0.361 | 1.48 | 1.02 - 2.16 | **0.042** | 0.71 | 0.45 - 1.12 | 0.143 |
| Prebiotics | 0.76 | 0.37 – 1.57 | 0.453 | 0.93 | 0.49 - 1.78 | 0.834 | 0.93 | 0.42 - 2.06 | 0.865 |

Results for each outcome variable were estimated using generalized linear regression with a Gamma distribution and log link and adjusted for covariates.
Fold Change, 95% Confidence Intervals (CI), and p-values are shown for each model.
Significant predictors (p<0.05) are indicated in bold.

**Supplementary Table 3A:**
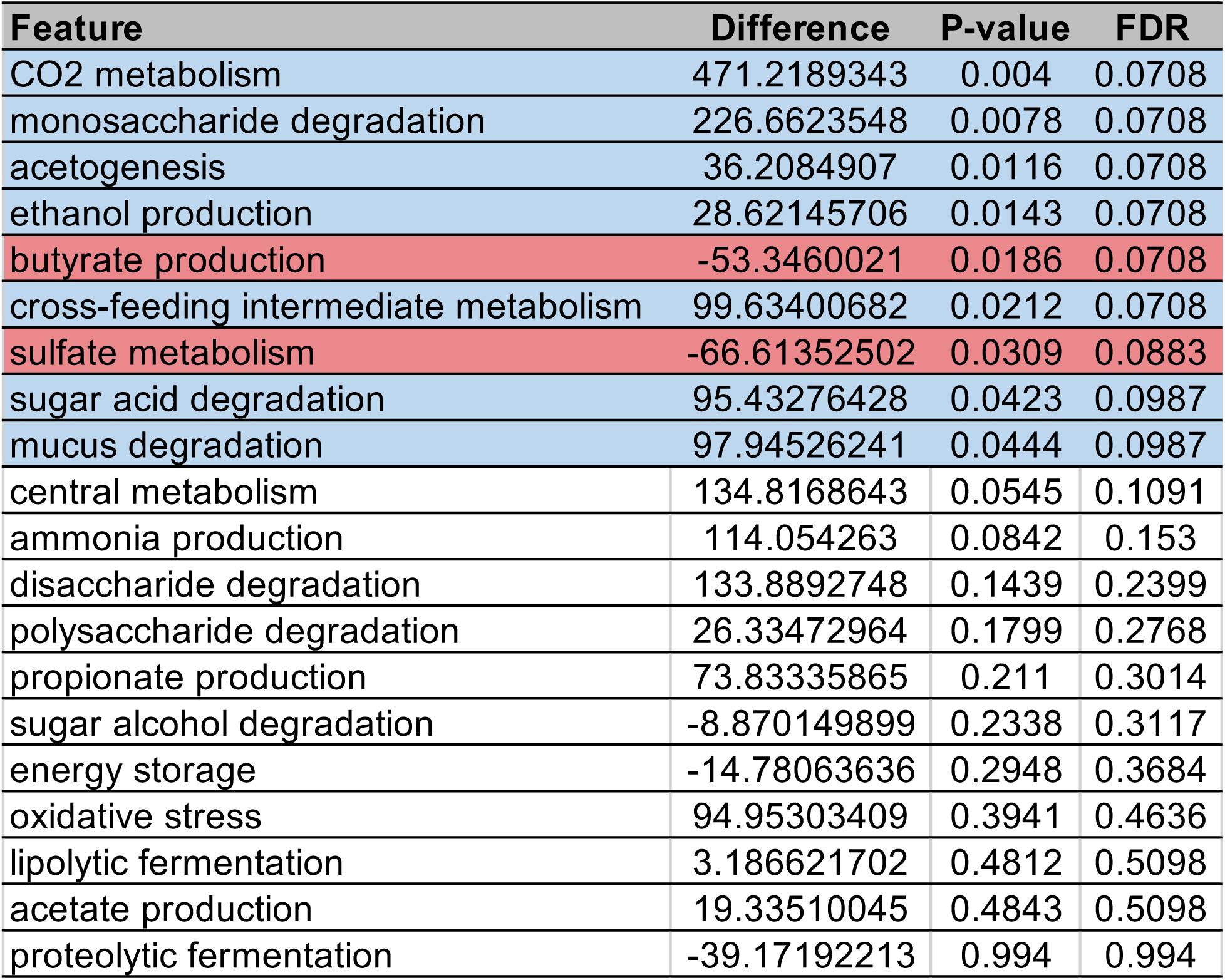
Functional metagenomic analysis of gut bacterial metabolic processes comparing ME/CFS cases vs. healthy controls using GOmixer analysis. Bacterial metabolic processes that differed between ME/CFS and controls (FDR adjusted p-value < 0.1) are highlighted in red (deficient in ME/CFS) and blue (enriched in ME/CFS). The mean difference, p-value, and FDR adjusted p-value are shown.

**Supplementary Table 3B.** Functional metagenomic analysis of gut bacterial metabolic modules comparing ME/CFS cases vs. healthy controls using GOmixer analysis. Bacterial metabolic modules that differed between ME/CFS and controls (FDR adjusted p-value < 0.1) are highlighted in red (deficient in ME/CFS) and blue (enriched in ME/CFS). The mean difference, p-value, and FDR adjusted p-value are shown.

| Feature | Difference | P-value | FDR |
| --- | --- | --- | --- |
| MF0119: lactate production | 107.35853 | 4.00E-04 | 0.0154 |
| MF0083: pyruvate dehydrogenase complex | 82.678087 | 4.00E-04 | 0.0154 |
| MF0060: ribose degradation | 40.029108 | 6.00E-04 | 0.0154 |
| MF0133: menaquinone production | 100.70313 | 0.0024 | 0.0449 |
| MF0132: superoxide reductase | -218.61741 | 0.0042 | 0.0531 |
| MF0014: glutamate degradation (4-aminobutanoate pathway) | 64.023843 | 0.0048 | 0.0531 |
| MF0055: xylose degradation | 61.010894 | 0.0051 | 0.0531 |
| MF0116: butyrate production via transferase | -48.018305 | 0.0056 | 0.0531 |
| MF0082: pentose phosphate pathway (oxidative branch) | 50.960008 | 0.0104 | 0.0718 |
| MF0085: pyruvate:formate lyase | 64.143101 | 0.0114 | 0.0718 |
| MF0054: arabinose degradation | 236.26021 | 0.0114 | 0.0718 |
| MF0093: homoacetogenesis | 36.208491 | 0.0116 | 0.0718 |
| MF0073: sorbitol degradation (dehydrogenase) | -37.037015 | 0.0123 | 0.0718 |
| MF0001: ethanol production (formate pathway) | 28.621457 | 0.0143 | 0.0736 |
| MF0131: superoxide dismutase | 241.32596 | 0.0145 | 0.0736 |
| MF0041: histidine degradation | 39.541857 | 0.0171 | 0.0814 |
| MF0052: chondroitin sulfate and dermatan sulfate degradation | 16.951528 | 0.0197 | 0.0829 |
| MF0108: glycerol degradation (dihydroxyacetone pathway) | -39.913774 | 0.0215 | 0.0829 |
| MF0086: TCA cycle | 22.568684 | 0.0218 | 0.0829 |
| MF0006: urea degradation | -17.960489 | 0.0218 | 0.0829 |
| MF0081: Glycolysis (pay-off phase) | -42.238444 | 0.0242 | 0.0877 |
| MF0100: Sulfate reduction (dissimilatory) | -34.755555 | 0.03 | 0.1035 |
| MF0102: mucin degradation | 67.000474 | 0.0331 | 0.1094 |
| MF0043: arginine degradation (agmatinase pathway) | -30.109059 | 0.0389 | 0.1231 |
| MF0071: D-galacturonate degradation | 46.380643 | 0.0434 | 0.129 |
| MF0023: methionine degradation (cysteine pathway) | -67.360619 | 0.0441 | 0.129 |
| MF0034: glutamine degradation (oxoglutarate pathway) | -16.249549 | 0.0636 | 0.1631 |
| MF0087: TCA cycle (Mycobacterium pathway) | 27.68447 | 0.0654 | 0.1631 |
| MF0032: glutamine degradation (ammonia pathway) | 51.234109 | 0.0654 | 0.1631 |
| MF0009: tryptophan degradation | 43.282852 | 0.0654 | 0.1631 |
| MF0091: beta-D-glucuronide and D-glucuronate degradation | 62.426313 | 0.0665 | 0.1631 |
| MF0035: arginine degradation (agmatine deiminase pathway) | -26.505721 | 0.0793 | 0.1756 |
| MF0016: glycine degradation | 39.622538 | 0.0806 | 0.1756 |
| MF0028: serine degradation | -82.123578 | 0.0833 | 0.1756 |
| MF0112: acetate to acetyl-CoA | 14.832126 | 0.0846 | 0.1756 |
| MF0110: glyoxylate bypass | 41.34142 | 0.0855 | 0.1756 |
| MF0048: lactose degradation | 97.925385 | 0.0855 | 0.1756 |
| MF0040: lysine degradation (cadaverine pathway) | 0 | 0.0878 | 0.1756 |
| MF0080: Glycolysis (preparatory phase) | -13.413274 | 0.1022 | 0.1992 |
| MF0088: TCA cycle (Helicobacter pathway) | 14.144595 | 0.1132 | 0.215 |
| MF0020: valine degradation | -16.192223 | 0.1232 | 0.2283 |
| MF0056: galactose degradation (Leloir pathway) | 26.195963 | 0.1281 | 0.2319 |
| MF0031: asparagine degradation | 77.929697 | 0.1412 | 0.2495 |
| MF0030: threonine degradation (formate pathway) | 15.321248 | 0.1509 | 0.2607 |
| MF0114: acetyl-CoA to crotonyl-CoA | -23.8441 | 0.1783 | 0.3011 |
| MF0065: pectin degradation - 5-dehydro-4-deoxy-D-glucuronate degradation | 22.481844 | 0.2047 | 0.3382 |
| MF0122: propionate production (succinate pathway) | 73.833359 | 0.211 | 0.3412 |
| MF0058: fructose degradation | -13.894422 | 0.2279 | 0.3609 |
| MF0130: peroxidase | 50.776936 | 0.2418 | 0.369 |
| MF0050: melibiose degradation | 32.928023 | 0.2428 | 0.369 |
| MF0022: isoleucine degradation | -10.119973 | 0.2902 | 0.426 |
| MF0066: glycogen metabolism | -14.780636 | 0.2948 | 0.426 |
| MF0005: acetylneuraminate and acetylmannosamine degradation | 9.4915474 | 0.2971 | 0.426 |
| MF0074: mannitol degradation | 6.1839954 | 0.337 | 0.4744 |
| MF0003: acetylglucosamine degradation | 25.612869 | 0.3655 | 0.5051 |
| MF0033: cysteine degradation (mercaptopyruvate pathway) | -12.304581 | 0.4327 | 0.5769 |
| MF0011: aspartate degradation (oxaloacetate pathway) | -24.609163 | 0.4327 | 0.5769 |
| MF0129: catalase | -0.9498511 | 0.4408 | 0.5777 |
| MF0117: butyrate production via kinase | -22.162235 | 0.4506 | 0.5804 |
| MF0092: kdo2-lipid A synthesis | -1.0948324 | 0.4704 | 0.59 |
| MF0063: fructan degradation | -19.456425 | 0.4812 | 0.59 |
| MF0113: acetyl-CoA to acetate | 19.3351 | 0.4843 | 0.59 |
| MF0090: pentose phosphate pathway (non-oxidative branch) | -14.773724 | 0.4891 | 0.59 |
| MF0062: starch degradation | -19.163231 | 0.4985 | 0.592 |
| MF0127: Succinate production | -3.2053077 | 0.5211 | 0.6093 |
| MF0018: proline degradation (glutamate pathway) | -1.2827993 | 0.5643 | 0.6498 |
| MF0079: bifidobacterium shunt | -18.576717 | 0.6464 | 0.7333 |
| MF0061: mannose degradation | 7.46386 | 0.6573 | 0.7346 |
| MF0109: glycerol degradation (glycerol kinase pathway) | 4.0745394 | 0.6902 | 0.7564 |
| MF0101: Sulfate reduction (assimilatory) | -7.0973628 | 0.6967 | 0.7564 |
| MF0012: aspartate degradation (fumarate pathway) | 7.7925417 | 0.7181 | 0.7687 |
| MF0059: rhamnose degradation | -4.5216527 | 0.8196 | 0.8651 |
| MF0046: sucrose degradation | 16.470197 | 0.8548 | 0.8899 |
| MF0029: threonine degradation (glycine pathway) | -13.554752 | 0.8844 | 0.9083 |
| MF0124: Fucose degradation | -4.9981401 | 0.966 | 0.9789 |
| MF0084: pyruvate:ferredoxin oxidoreductase | -2.1354248 | 0.982 | 0.982 |

**Supplementary Table 3C.** Functional metagenomic analysis of gut bacterial metabolic modules comparing ME/CFS cases vs. healthy controls using GOmixer analysis with gut-brain specific modules. Bacterial gut-brain metabolic modules that differed between ME/CFS and controls (FDR adjusted p-value < 0.1) are highlighted in red (deficient in ME/CFS) and blue (enriched in ME/CFS). The mean difference, p-value, and FDR adjusted p-value are shown.

| Feature | Difference | P-value | FDR |
| --- | --- | --- | --- |
| MGB029: ClpB (ATP-dependent chaperone protein) | -65.07429824 | 0.001 | 0.0156 |
| MGB040: Menaquinone synthesis (vitamin K2) I | 47.56075313 | 0.0016 | 0.0156 |
| MGB022: GABA synthesis III | 64.02384314 | 0.0048 | 0.0279 |
| MGB053: Butyrate synthesis II | -48.0183053 | 0.0056 | 0.0279 |
| MGB036: S-Adenosylmethionine (SAM) synthesis | -136.753975 | 0.0082 | 0.0327 |
| MGB034: Isovaleric acid synthesis I (KADH pathway) | 29.33347714 | 0.0107 | 0.0357 |
| MGB007: Glutamate synthesis II | -16.24954924 | 0.0636 | 0.1636 |
| MGB049: Tryptophan degradation | 43.28285214 | 0.0654 | 0.1636 |
| MGB047: Acetate degradation | 14.83212637 | 0.0846 | 0.1832 |
| MGB037: Inositol synthesis | 52.01371852 | 0.0916 | 0.1832 |
| MGB038: Inositol degradation | -19.22224684 | 0.1184 | 0.2152 |
| MGB043: Acetate synthesis I | 15.84392198 | 0.3602 | 0.581 |
| MGB055: Propionate synthesis III | 51.4889175 | 0.3872 | 0.581 |
| MGB005: Tryptophan synthesis | 10.0869699 | 0.4067 | 0.581 |
| MGB052: Butyrate synthesis I | -22.16223543 | 0.4506 | 0.6007 |
| MGB032: Quinolinic acid synthesis | -6.749680792 | 0.5049 | 0.6312 |
| MGB033: Quinolinic acid degradation | 4.261764131 | 0.566 | 0.6658 |
| MGB031: 17-beta-Estradiol degradation | 20.0768445 | 0.6268 | 0.6964 |
| MGB006: Glutamate synthesis I | -54.06773541 | 0.6718 | 0.7072 |
| MGB015: p-Cresol synthesis | -6.858126161 | 0.8118 | 0.8118 |

**Supplementary Table 4A:**
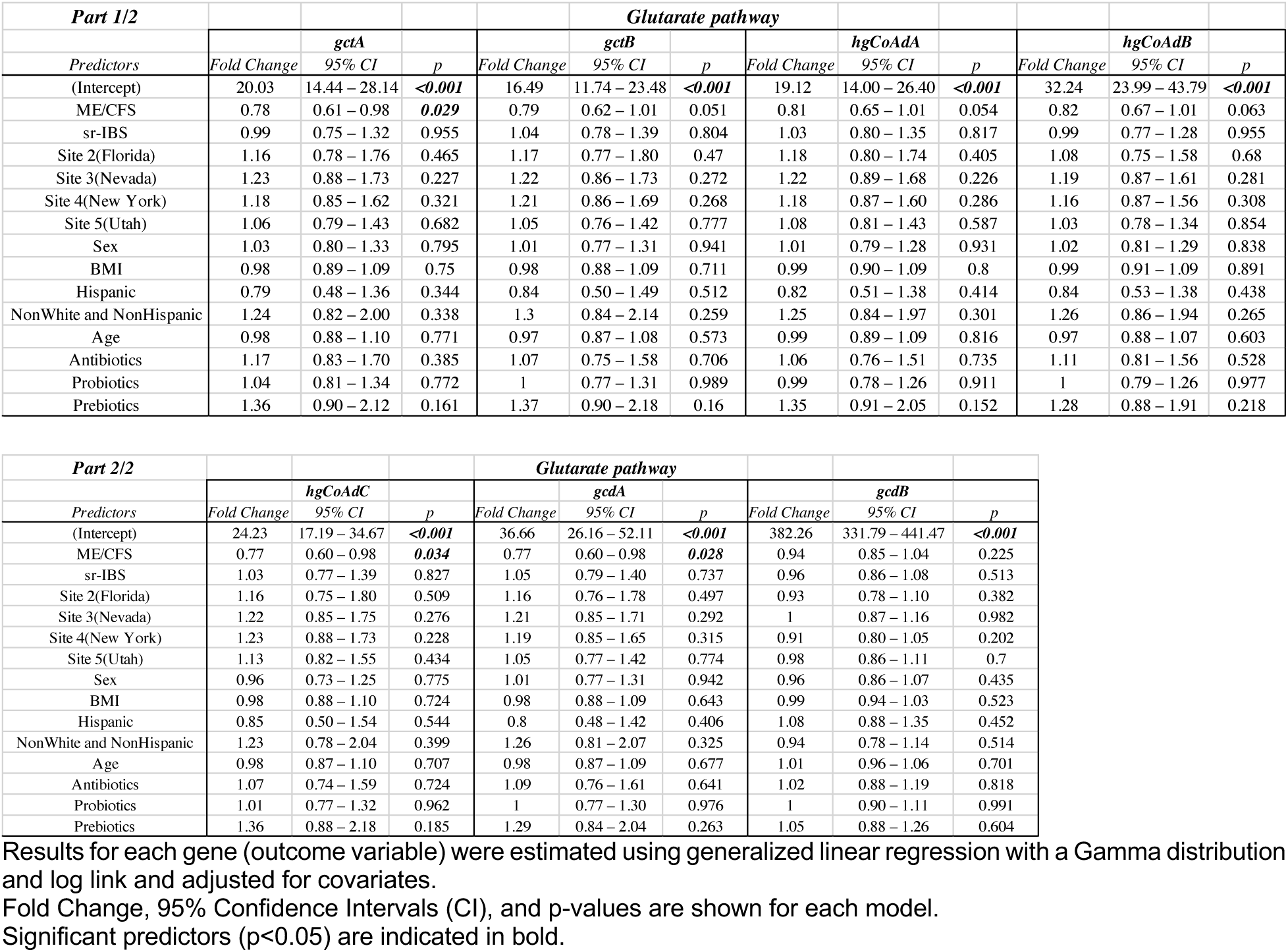
Generalized linear regression showing the relationship between ME/CFS status and the metagenomic counts (CPM) of genes along the Glutarate pathway of butyrate production.

**Supplementary Table 4B.** Generalized linear regression showing the relationship between ME/CFS status and the metagenomic counts (CPM) of genes along the Lysine pathway of butyrate production.

| Part 1/2 |  |  |  |  | Lysine pathway |  |  |  |  |  |  |  |  |
| --- | --- | --- | --- | --- | --- | --- | --- | --- | --- | --- | --- | --- | --- |
|  | Predictors | Fold Change | kamA<br>95% CI | p | Fold Change | kamD<br>95% CI | p | Fold Change | kamE<br>95% CI | p | Fold Change | kdd<br>95% CI | p |
| (Intercept) |  | 93.61 | 63.67 – 140.07 | <0.001 | 121.56 | 84.36 – 178.01 | <0.001 | 62.02 | 42.15 – 92.87 | <0.001 | 105.85 | 77.68 – 145.95 | <0.001 |
| ME/CFS |  | 0.81 | 0.63 – 1.06 | 0.129 | 0.82 | 0.64 – 1.05 | 0.122 | 0.81 | 0.62 – 1.06 | 0.129 | 0.88 | 0.71 – 1.09 | 0.252 |
| sr-IBS |  | 1.01 | 0.73 – 1.41 | 0.939 | 1 | 0.73 – 1.37 | 0.992 | 0.99 | 0.71 – 1.38 | 0.943 | 1.13 | 0.87 – 1.47 | 0.37 |
| Site 2(Florida) |  | 0.75 | 0.47 – 1.21 | 0.228 | 0.73 | 0.47 – 1.16 | 0.174 | 0.72 | 0.45 – 1.17 | 0.172 | 0.86 | 0.59 – 1.27 | 0.443 |
| Site 3(Nevada) |  | 0.97 | 0.65 – 1.45 | 0.881 | 0.98 | 0.67 – 1.43 | 0.901 | 0.95 | 0.63 – 1.42 | 0.803 | 1 | 0.72 – 1.38 | 0.992 |
| Site 4(New York) |  | 0.79 | 0.54 – 1.16 | 0.224 | 0.8 | 0.56 – 1.16 | 0.234 | 0.8 | 0.54 – 1.17 | 0.241 | 0.91 | 0.67 – 1.23 | 0.523 |
| Site 5(Utah) |  | 0.97 | 0.67 – 1.37 | 0.85 | 0.98 | 0.70 – 1.37 | 0.907 | 0.97 | 0.68 – 1.38 | 0.86 | 1.01 | 0.75 – 1.34 | 0.963 |
| Sex |  | 1.05 | 0.77 – 1.40 | 0.762 | 1.04 | 0.78 – 1.37 | 0.79 | 1.05 | 0.78 – 1.41 | 0.724 | 1.04 | 0.82 – 1.32 | 0.73 |
| BMI |  | 1.07 | 0.94 – 1.21 | 0.296 | 1.09 | 0.97 – 1.22 | 0.148 | 1.08 | 0.96 – 1.23 | 0.188 | 1.07 | 0.97 – 1.18 | 0.184 |
| Hispanic |  | 1.9 | 1.07 – 3.62 | 0.031 | 1.9 | 1.10 – 3.50 | 0.024 | 1.99 | 1.11 – 3.82 | 0.021 | 1.74 | 1.09 – 2.91 | 0.022 |
| NonWhite and NonHispanic |  | 0.5 | 0.31 – 0.89 | 0.011 | 0.53 | 0.33 – 0.92 | 0.014 | 0.51 | 0.31 – 0.90 | 0.012 | 0.55 | 0.37 – 0.86 | 0.006 |
| Age |  | 1.12 | 0.99 – 1.28 | 0.074 | 1.12 | 0.99 – 1.26 | 0.074 | 1.14 | 1.00 – 1.29 | 0.048 | 1.07 | 0.96 – 1.18 | 0.226 |
| Antibiotics |  | 0.73 | 0.48 – 1.14 | 0.132 | 0.78 | 0.53 – 1.19 | 0.215 | 0.73 | 0.48 – 1.14 | 0.133 | 0.82 | 0.59 – 1.17 | 0.246 |
| Probiotics |  | 0.97 | 0.73 – 1.31 | 0.849 | 0.97 | 0.74 – 1.30 | 0.841 | 0.97 | 0.72 – 1.32 | 0.839 | 0.99 | 0.78 – 1.26 | 0.932 |
| Prebiotics |  | 1.14 | 0.71 – 1.92 | 0.601 | 1.2 | 0.77 – 1.98 | 0.442 | 1.18 | 0.74 – 2.00 | 0.509 | 1.17 | 0.79 – 1.78 | 0.443 |

| *Part 2/2* | *Lysine pathway* |  |  |  |  |  |  |  |  |  |  |  |
| --- | --- | --- | --- | --- | --- | --- | --- | --- | --- | --- | --- | --- |
| *Predictors* | *kce* |  |  | *kal* |  |  | *atoA* |  |  | *atoD* |  |  |
|  | *Fold Change* | *95% CI* | *p* | *Fold Change* | *95% CI* | *p* | *Fold Change* | *95% CI* | *p* | *Fold Change* | *95% CI* | *p* |
| (Intercept) | 65.59 | 46.61 – 93.61 | **<0.001** | 32.05 | 21.86 – 47.81 | **<0.001** | 51.89 | 35.54 – 77.07 | **<0.001** | 49.06 | 32.62 – 75.27 | **<0.001** |
| ME/CFS | 0.91 | 0.72 – 1.15 | 0.439 | 0.86 | 0.66 – 1.11 | 0.254 | 0.76 | 0.59 – 0.98 | **0.042** | 0.71 | 0.54 – 0.94 | **0.02** |
| sr-IBS | 1.09 | 0.82 – 1.46 | 0.557 | 1 | 0.73 – 1.39 | 0.992 | 0.98 | 0.71 – 1.36 | 0.883 | 0.98 | 0.70 – 1.41 | 0.93 |
| Site 2(Florida) | 0.79 | 0.52 – 1.21 | 0.272 | 0.72 | 0.45 – 1.16 | 0.166 | 0.78 | 0.49 – 1.25 | 0.289 | 0.77 | 0.46 – 1.29 | 0.302 |
| Site 3(Nevada) | 1 | 0.70 – 1.43 | 0.985 | 0.97 | 0.65 – 1.44 | 0.868 | 0.91 | 0.61 – 1.35 | 0.625 | 0.89 | 0.58 – 1.36 | 0.583 |
| Site 4(New York) | 0.89 | 0.64 – 1.26 | 0.514 | 0.83 | 0.57 – 1.21 | 0.327 | 0.78 | 0.54 – 1.15 | 0.201 | 0.79 | 0.53 – 1.19 | 0.255 |
| Site 5(Utah) | 0.99 | 0.72 – 1.35 | 0.944 | 0.96 | 0.68 – 1.36 | 0.837 | 0.96 | 0.68 – 1.36 | 0.839 | 0.99 | 0.68 – 1.44 | 0.965 |
| Sex | 1.11 | 0.85 – 1.45 | 0.424 | 1.04 | 0.77 – 1.39 | 0.804 | 1.02 | 0.76 – 1.37 | 0.881 | 0.97 | 0.70 – 1.32 | 0.837 |
| BMI | 1.07 | 0.96 – 1.19 | 0.244 | 1.07 | 0.95 – 1.21 | 0.25 | 1.09 | 0.97 – 1.23 | 0.159 | 1.1 | 0.97 – 1.26 | 0.135 |
| Hispanic | 1.99 | 1.19 – 3.52 | **0.009** | 1.87 | 1.05 – 3.56 | **0.034** | 1.86 | 1.06 – 3.51 | **0.034** | 1.8 | 0.98 – 3.59 | 0.063 |
| NonWhite and NonHispanic | 0.48 | 0.31 – 0.79 | **0.002** | 0.53 | 0.32 – 0.93 | **0.016** | 0.47 | 0.29 – 0.83 | **0.005** | 0.54 | 0.32 – 1.00 | **0.033** |
| Age | 1.06 | 0.95 – 1.19 | 0.279 | 1.11 | 0.98 – 1.26 | 0.099 | 1.13 | 1.00 – 1.28 | 0.056 | 1.14 | 0.99 – 1.30 | 0.061 |
| Antibiotics | 0.84 | 0.59 – 1.24 | 0.361 | 0.78 | 0.52 – 1.21 | 0.233 | 0.74 | 0.49 – 1.15 | 0.149 | 0.72 | 0.46 – 1.16 | 0.145 |
| Probiotics | 1.03 | 0.80 – 1.35 | 0.808 | 0.99 | 0.74 – 1.34 | 0.97 | 1.01 | 0.76 – 1.35 | 0.963 | 1.02 | 0.74 – 1.40 | 0.919 |
| Prebiotics | 1.22 | 0.80 – 1.94 | 0.381 | 1.13 | 0.71 – 1.90 | 0.622 | 1.18 | 0.74 – 1.97 | 0.514 | 1.31 | 0.79 – 2.30 | 0.317 |

Results for each gene (outcome variable) were estimated using generalized linear regression with a Gamma distribution and log link and adjusted for covariates.
Fold Change, 95% Confidence Intervals (CI), and p-values are shown for each model.
Significant predictors (p<0.05) are indicated in bold.

**Supplementary Table 4C.** Generalized linear regression showing the relationship between ME/CFS status and the metagenomic counts (CPM) of genes along the 4-Aminobutyrate pathway of butyrate production.

|  | *4-Aminobutyrate pathway* |  |  |  |  |  |  |  |  |
| --- | --- | --- | --- | --- | --- | --- | --- | --- | --- |
|  | *abfH* |  |  | *4hbt* |  |  | *abfD* |  |  |
| *Predictors* | *Fold Change* | *95% CI* | *p* | *Fold Change* | *95% CI* | *p* | *Fold Change* | *95% CI* | *p* |
| (Intercept) | 47.23 | 34.49 – 65.52 | **<0.001** | 44.5 | 33.07 – 60.55 | **<0.001** | 71.56 | 52.98 – 97.86 | **<0.001** |
| ME/CFS | 0.96 | 0.76 – 1.21 | 0.744 | 1.02 | 0.83 – 1.26 | 0.869 | 1.01 | 0.80 – 1.26 | 0.957 |
| sr-IBS | 1.29 | 0.98 – 1.70 | 0.064 | 1.24 | 0.97 – 1.61 | 0.083 | 1.11 | 0.85 – 1.45 | 0.449 |
| Site 2(Florida) | 0.89 | 0.60 – 1.33 | 0.572 | 0.85 | 0.59 – 1.23 | 0.38 | 0.98 | 0.67 – 1.44 | 0.921 |
| Site 3(Nevada) | 0.97 | 0.70 – 1.34 | 0.857 | 0.9 | 0.66 – 1.23 | 0.502 | 0.97 | 0.71 – 1.33 | 0.866 |
| Site 4(New York) | 1.08 | 0.78 – 1.48 | 0.651 | 0.92 | 0.69 – 1.23 | 0.574 | 0.96 | 0.71 – 1.31 | 0.817 |
| Site 5(Utah) | 0.99 | 0.73 – 1.32 | 0.935 | 1 | 0.76 – 1.31 | 0.983 | 0.93 | 0.70 – 1.24 | 0.637 |
| Sex | 1.04 | 0.81 – 1.32 | 0.775 | 1.09 | 0.86 – 1.37 | 0.482 | 1.1 | 0.87 – 1.39 | 0.435 |
| BMI | 1 | 0.90 – 1.11 | 1.00 | 1.03 | 0.94 – 1.13 | 0.553 | 0.96 | 0.87 – 1.06 | 0.379 |
| Hispanic | 1.43 | 0.89 – 2.41 | 0.154 | 1.33 | 0.86 – 2.17 | 0.209 | 1.14 | 0.71 – 1.90 | 0.596 |
| NonWhite and NonHispanic | 0.89 | 0.58 – 1.42 | 0.594 | 0.63 | 0.43 – 0.96 | **0.024** | 0.88 | 0.58 – 1.38 | 0.541 |
| Age | 1.01 | 0.91 – 1.12 | 0.886 | 1.01 | 0.92 – 1.12 | 0.777 | 1.04 | 0.94 – 1.16 | 0.449 |
| Antibiotics | 0.93 | 0.67 – 1.34 | 0.697 | 0.9 | 0.65 – 1.25 | 0.502 | 0.83 | 0.59 – 1.18 | 0.27 |
| Probiotics | 1.22 | 0.96 – 1.56 | 0.11 | 1.09 | 0.87 – 1.37 | 0.458 | 1.15 | 0.91 – 1.47 | 0.236 |
| Prebiotics | 1.02 | 0.68 – 1.58 | 0.926 | 1.23 | 0.84 – 1.84 | 0.3 | 0.99 | 0.66 – 1.51 | 0.949 |

Results for each gene (outcome variable) were estimated using generalized linear regression with a Gamma distribution and log link and adjusted for covariates.
Fold Change, 95% Confidence Intervals (CI), and p-values are shown for each model.
Significant predictors (p<0.05) are indicated in bold.

**Supplementary Table 5A:**
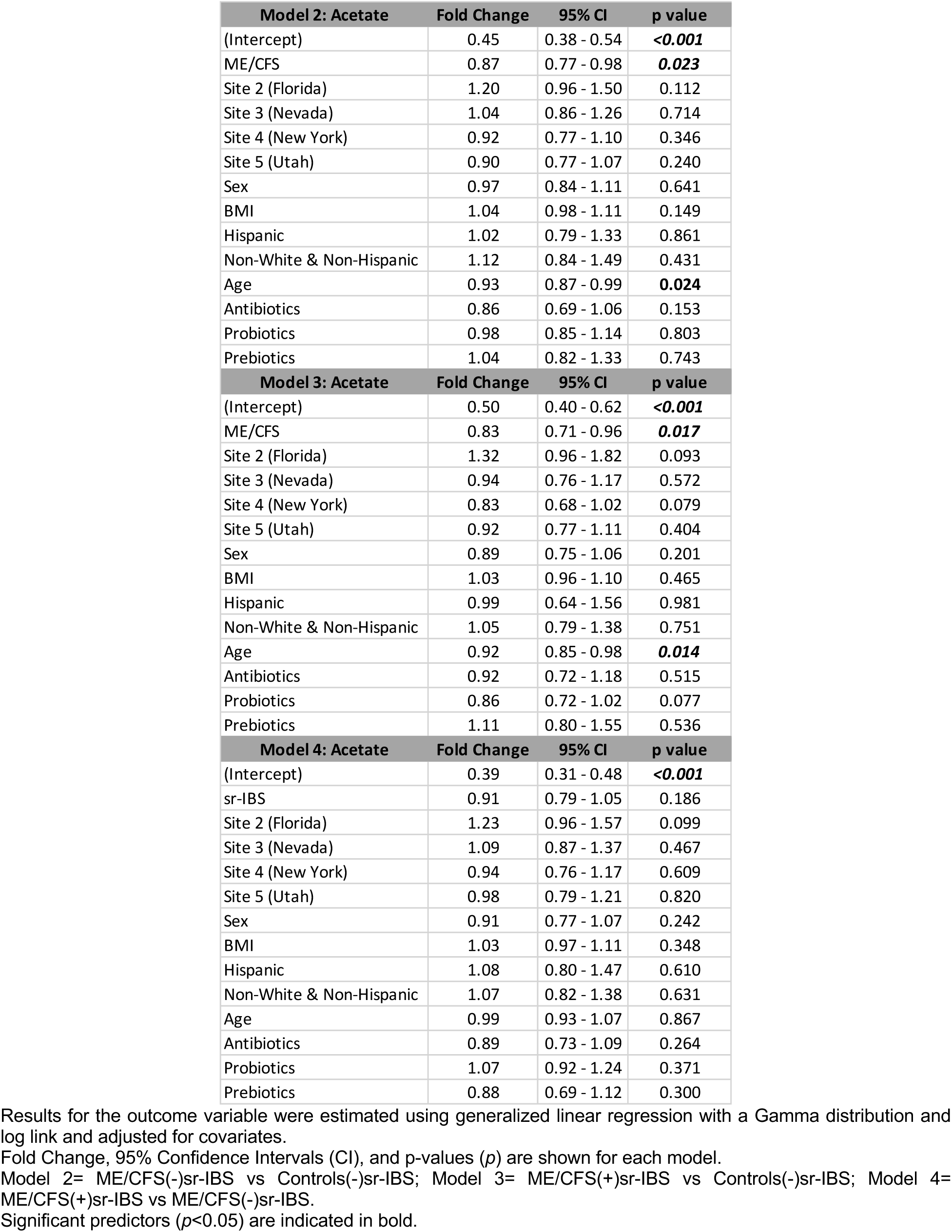
Generalized linear regression showing the relationship between ME/CFS status and fecal acetate concentrations.

| Model 2: Acetate | Fold Change | 95% CI | p value |
| --- | --- | --- | --- |
| (Intercept) | 0.45 | 0.38 - 0.54 | **<0.001** |
| ME/CFS | 0.87 | 0.77 - 0.98 | **0.023** |
| Site 2 (Florida) | 1.20 | 0.96 - 1.50 | 0.112 |
| Site 3 (Nevada) | 1.04 | 0.86 - 1.26 | 0.714 |
| Site 4 (New York) | 0.92 | 0.77 - 1.10 | 0.346 |
| Site 5 (Utah) | 0.90 | 0.77 - 1.07 | 0.240 |
| Sex | 0.97 | 0.84 - 1.11 | 0.641 |
| BMI | 1.04 | 0.98 - 1.11 | 0.149 |
| Hispanic | 1.02 | 0.79 - 1.33 | 0.861 |
| Non-White & Non-Hispanic | 1.12 | 0.84 - 1.49 | 0.431 |
| Age | 0.93 | 0.87 - 0.99 | **0.024** |
| Antibiotics | 0.86 | 0.69 - 1.06 | 0.153 |
| Probiotics | 0.98 | 0.85 - 1.14 | 0.803 |
| Prebiotics | 1.04 | 0.82 - 1.33 | 0.743 |
| Model 3: Acetate | Fold Change | 95% CI | p value |
| (Intercept) | 0.50 | 0.40 - 0.62 | **<0.001** |
| ME/CFS | 0.83 | 0.71 - 0.96 | **0.017** |
| Site 2 (Florida) | 1.32 | 0.96 - 1.82 | 0.093 |
| Site 3 (Nevada) | 0.94 | 0.76 - 1.17 | 0.572 |
| Site 4 (New York) | 0.83 | 0.68 - 1.02 | 0.079 |
| Site 5 (Utah) | 0.92 | 0.77 - 1.11 | 0.404 |
| Sex | 0.89 | 0.75 - 1.06 | 0.201 |
| BMI | 1.03 | 0.96 - 1.10 | 0.465 |
| Hispanic | 0.99 | 0.64 - 1.56 | 0.981 |
| Non-White & Non-Hispanic | 1.05 | 0.79 - 1.38 | 0.751 |
| Age | 0.92 | 0.85 - 0.98 | **0.014** |
| Antibiotics | 0.92 | 0.72 - 1.18 | 0.515 |
| Probiotics | 0.86 | 0.72 - 1.02 | 0.077 |
| Prebiotics | 1.11 | 0.80 - 1.55 | 0.536 |
| Model 4: Acetate | Fold Change | 95% CI | p value |
| (Intercept) | 0.39 | 0.31 - 0.48 | **<0.001** |
| sr-IBS | 0.91 | 0.79 - 1.05 | 0.186 |
| Site 2 (Florida) | 1.23 | 0.96 - 1.57 | 0.099 |
| Site 3 (Nevada) | 1.09 | 0.87 - 1.37 | 0.467 |
| Site 4 (New York) | 0.94 | 0.76 - 1.17 | 0.609 |
| Site 5 (Utah) | 0.98 | 0.79 - 1.21 | 0.820 |
| Sex | 0.91 | 0.77 - 1.07 | 0.242 |
| BMI | 1.03 | 0.97 - 1.11 | 0.348 |
| Hispanic | 1.08 | 0.80 - 1.47 | 0.610 |
| Non-White & Non-Hispanic | 1.07 | 0.82 - 1.38 | 0.631 |
| Age | 0.99 | 0.93 - 1.07 | 0.867 |
| Antibiotics | 0.89 | 0.73 - 1.09 | 0.264 |
| Probiotics | 1.07 | 0.92 - 1.24 | 0.371 |
| Prebiotics | 0.88 | 0.69 - 1.12 | 0.300 |

Results for the outcome variable were estimated using generalized linear regression with a Gamma distribution and log link and adjusted for covariates.
Fold Change, 95% Confidence Intervals (CI), and p-values (p) are shown for each model.
Model 2= ME/CFS(-)sr-IBS vs Controls(-)sr-IBS; Model 3= ME/CFS(+)sr-IBS vs Controls(-)sr-IBS; Model 4= ME/CFS(+)sr-IBS vs ME/CFS(-)sr-IBS.
Significant predictors (p<0.05) are indicated in bold.

**Supplementary Table 5B.** Generalized linear regression showing the relationship between ME/CFS status and fecal butyrate concentrations.

| Model 2: Butyrate | Fold Change | 95% CI | p value |
| --- | --- | --- | --- |
| (Intercept) | 0.36 | 0.26 - 0.50 | **<0.001** |
| ME/CFS | 0.60 | 0.48 - 0.75 | **<0.001** |
| Site 2 (Florida) | 1.15 | 0.76 - 1.75 | 0.501 |
| Site 3 (Nevada) | 1.03 | 0.72 - 1.47 | 0.863 |
| Site 4 (New York) | 0.83 | 0.59 - 1.15 | 0.257 |
| Site 5 (Utah) | 1.01 | 0.74 - 1.38 | 0.948 |
| Sex | 0.74 | 0.57 - 0.97 | **0.029** |
| BMI | 1.03 | 0.92 - 1.14 | 0.643 |
| Hispanic | 1.44 | 0.89 - 2.35 | 0.142 |
| Non-White & Non-Hispanic | 1.41 | 0.83 - 2.39 | 0.212 |
| Age | 0.93 | 0.83 - 1.04 | 0.197 |
| Antibiotics | 0.85 | 0.58 - 1.26 | 0.433 |
| Probiotics | 1.08 | 0.82 - 1.42 | 0.586 |
| Prebiotics | 1.08 | 0.69 - 1.71 | 0.731 |
| Model 3: Butyrate | Fold Change | 95% CI | p value |
| (Intercept) | 0.35 | 0.24 - 0.52 | **<0.001** |
| ME/CFS | 0.56 | 0.42 - 0.73 | **<0.001** |
| Site 2 (Florida) | 1.11 | 0.62 - 2.00 | 0.715 |
| Site 3 (Nevada) | 1.06 | 0.72 - 1.58 | 0.760 |
| Site 4 (New York) | 0.76 | 0.53 - 1.10 | 0.150 |
| Site 5 (Utah) | 1.06 | 0.75 - 1.48 | 0.753 |
| Sex | 0.75 | 0.55 - 1.03 | 0.082 |
| BMI | 1.01 | 0.89 - 1.14 | 0.866 |
| Hispanic | 1.67 | 0.74 - 3.75 | 0.219 |
| Non-White & Non-Hispanic | 1.11 | 0.67 - 1.84 | 0.688 |
| Age | 0.89 | 0.78 - 1.01 | 0.071 |
| Antibiotics | 1.02 | 0.65 - 1.59 | 0.929 |
| Probiotics | 0.88 | 0.65 - 1.20 | 0.434 |
| Prebiotics | 1.36 | 0.75 - 2.49 | 0.317 |
| Model 4: Butyrate | Fold Change | 95% CI | p value |
| (Intercept) | 0.21 | 0.14 - 0.33 | **<0.001** |
| sr-IBS | 0.88 | 0.66 - 1.17 | 0.379 |
| Site 2 (Florida) | 1.31 | 0.80 - 2.14 | 0.287 |
| Site 3 (Nevada) | 1.25 | 0.79 - 1.99 | 0.342 |
| Site 4 (New York) | 0.86 | 0.56 - 1.34 | 0.516 |
| Site 5 (Utah) | 0.97 | 0.63 - 1.50 | 0.892 |
| Sex | 0.72 | 0.52 - 1.00 | 0.051 |
| BMI | 0.97 | 0.84 - 1.11 | 0.631 |
| Hispanic | 1.43 | 0.77 - 2.65 | 0.261 |
| Non-White & Non-Hispanic | 1.13 | 0.67 - 1.91 | 0.641 |
| Age | 0.94 | 0.82 - 1.09 | 0.427 |
| Antibiotics | 0.93 | 0.63 - 1.39 | 0.739 |
| Probiotics | 1.11 | 0.83 - 1.50 | 0.474 |
| Prebiotics | 0.81 | 0.50 - 1.31 | 0.395 |

Results for the outcome variable were estimated using generalized linear regression with a Gamma distribution and log link and adjusted for covariates.
Fold Change, 95% Confidence Intervals (CI), and p-values (p) are shown for each model.
Model 2= ME/CFS(-)sr-IBS vs Controls(-)sr-IBS; Model 3= ME/CFS(+)sr-IBS vs Controls(-)sr-IBS; Model 4= ME/CFS(+)sr-IBS vs ME/CFS(-)sr-IBS.
Significant predictors ( $p < 0.05$ ) are indicated in bold.

**Supplementary Table 5C.**
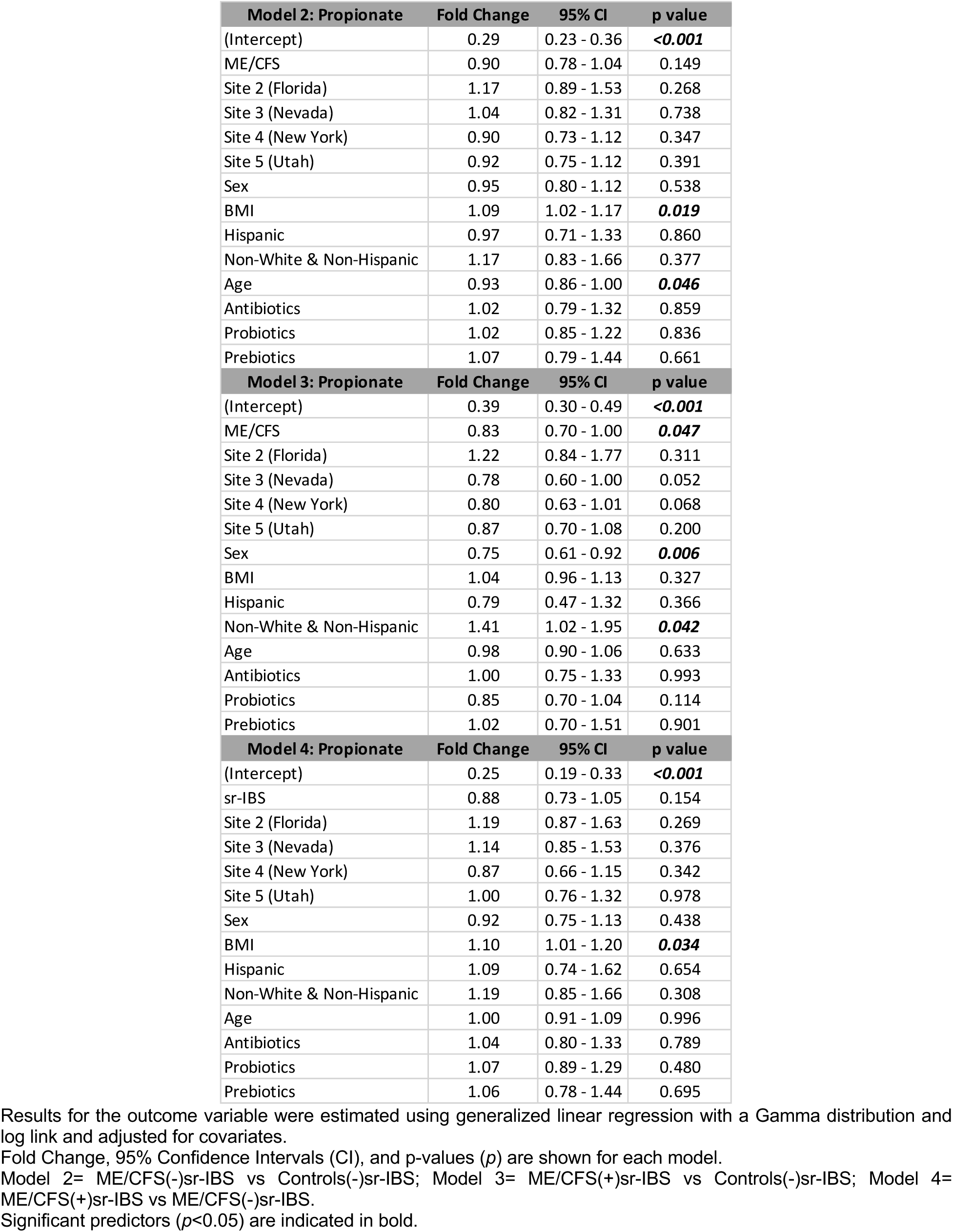
Generalized linear regression showing the relationship between ME/CFS status and fecal propionate concentrations.

**Supplementary Table 6:** Comparison of MFI dimension scores between groups.

| Fatigue Dimensions | ME/CFS (n=106) | Healthy controls (n=91) | p-value | ME/CFS (-) sr-IBS (n=71) | ME/CFS (+) sr-IBS (n=35) | p-value |
| --- | --- | --- | --- | --- | --- | --- |
| General fatigue (Median) | 91.67 | 16.67 | p < 0.0001 | 91.67 | 95.83 | 0.9660 |
| Physical fatigue (Median) | 87.50 | 12.50 | p < 0.0001 | 83.33 | 91.67 | 0.0878 |
| Mental fatigue (Median) | 62.50 | 12.50 | p < 0.0001 | 58.33 | 66.67 | 0.2942 |
| Reduced activity (Median) | 79.17 | 4.17 | p < 0.0001 | 75.00 | 87.50 | **0.0189** |
| Reduced motivation (Median) | 45.83 | 12.50 | p < 0.0001 | 45.83 | 50.00 | 0.5168 |

p-values based on two-tailed Mann-Whitney U test

